# SARS-CoV-2 infection detection efficiency in Denmark before major vaccine impacts

**DOI:** 10.64898/2026.10.16.26364986

**Authors:** Morten N. Westfall, Niels Høiby, Steen Rasmussen, Hans-Joachim Ziock

**Affiliations:** Center for Økonomi, Region Hovedstaden; Department of Clinical Microbiology, Rigshospitalet, Denmark; Institute of Immunology and Microbiology, University of Copenhagen, Denmark; University of Southern Denmark; Santa Fe Institute, NM U.S.A; European Centre for Living Technology (ECLT), Venice, Italy; Los Alamos, NM U.S.A

## Abstract

To deal with the COVID-19 pandemic, Denmark implemented an aggressive program aimed at controlling the pandemic which included both testing and contact tracing programs. The large amount of data collected in Denmark on the pandemic and its integrated nature made it possible to investigate the effectiveness of the testing and contact tracing effort. The results of that investigation and a discussion of the effectiveness and limitations of the different control efforts are reported herein, as are lessons learned concerning possible improvements for the future. The analysis is limited to the period from the start of the COVID-19 pandemic in Denmark (late Feb. 2020) into May 2021, at which time the impact of vaccination intervention was already playing a role with ∼25% of the Danish population at least partially vaccinated. From the available data sets we estimate the national Infection Detection Efficiency (*IDE*) starting around July 1, 2020, the time at which the national infection detection program started to be implemented in a more robust form. The results rely on daily COVID-19 hospital admissions, PCR test results, and antibody prevalence values for the Danish population. We find that for the long-term average, the combined Danish national testing and contact tracing program identified on average about 55 to 60% of the infected individuals during the period from Sept. 1, 2020, into early May 2021. However, higher infection detection efficiencies were achieved in localized areas for short time intervals. Our investigations further indicate that the Danish testing and contact tracing program, which was among the most ambitious in the world, could have had a better balance between voluntary probing on the one hand and focused contact tracing and targeted testing on the other hand. As typically implemented, it proved to be largely ineffective in controlling the pandemic. However, in at least one case when it was implemented in an enhanced and focused mode, it was able to bring a large local outbreak that dominated the country’s cases under control. We have compiled a list of lessons learned regarding the use of contact tracing procedures to control potential future pandemics before vaccinations become available.

## 1. Introduction

Denmark implemented an aggressive program of actions to try to control the COVID-19 pandemic within its borders. These involved shutdowns of its society, building a vigorous testing program, implementing a large-scale contact tracing/testing program, vaccination of its population, etc. Herein we seek to evaluate the true effectiveness of the actions taken to limit/control the pandemic prior to the vaccinations becoming widespread.

Monitoring the true scale of the COVID-19 pandemic in real time is difficult as the infections have a wide range of manifestations; see for example Ref. [1]. Which ones are exhibited depend on the traits of the individuals who are infected. The manifestations range from no symptoms whatsoever, all the way to hospitalization and even death. The main difficulty in trying to evaluate the effectiveness of actions taken to limit/control the pandemic is the inability to actually know the scale of the pandemic due to the significant fraction of the pandemic that is carried and spread by people who have become contagious but do not yet show symptoms or those that remain asymptomatic throughout the course of their infection. Furthermore, unless an unbiased sampling of the general population for current and/or past COVID-19 infections can be established, it is impossible to ascertain the size of the asymptomatic fraction. This latter issue was addressed in Denmark by testing randomized samples of the Danish population at different times for past COVID-19 infections via COVID-19 antibodies while also questioning the participants about having had symptoms consistent with a COVID-19 infection. The results of those studies indicated that 41% of those with antibodies for COVID-19 had symptom-free COVID-19 infections [2]. Such a high percentage highlights the difficulty in counting the true daily number of new cases, let alone finding and isolating all those infected as a means of containing the COVID-19 pandemic. This was especially true early in the pandemic when those mildly infected would assume that they had a common cold and continue to mingle with the population at large. Furthermore, early on in the pandemic the availability of tests to confirm COVID-19 infections were either non-existent or extremely limited. Simply put, tests were unavailable in the numbers needed to deal with the scope of the outbreaks. The situation improved as adequate testing resources became available to deal with the number of symptomatic cases and improved even more as the testing capability further increased to allow testing of people identified by contact tracing. However, finding all current cases of COVID-19 remained difficult due to the significant fraction of those infected that never displayed any symptoms and/or were missed by contact tracing.

As a simple counting of positive COVID-19 PCR test results does not reveal the scope of the COVID-19 pandemic in Denmark (especially when testing capacity was insufficient), another method had to be found to provide a daily tracking of the COVID-19 pandemic in Denmark. We chose the daily number of COVID-19 related hospital admissions to do this. COVID-19 hospital admission decisions (admissions longer than 12 hours) are made by medical professionals and thus are fairly standardized and should be representative of the scale of the COVID-19 pandemic as a function of time. Of course, the number of hospital admissions had to be scaled to the actual number of COVID-19 cases. This was done by integrating the number of hospital admissions through a date corresponding to a COVID-19 antibody prevalence measurement representative of the Danish population and then using the ratio of those two quantities as the scaling factor. COVID-19 antibody prevalence values provide an integral number of COVID-19 cases as the antibodies are long-lived enough, given the high sensitivity of the antibody tests used, to remain detectable throughout the period studied. More details on this are given in Section 2 below. Concerning the use of hospital admissions, no better stand-in representative of the actual daily number of new COVID-19 cases could be identified. There are however a number of caveats concerning the use of the hospital admission values that are discussed in Section 3. As it turns out, adjustments to the raw hospital admission values are needed.

With a value in hand that is representative of the actual daily number of new COVID-19 cases, that value can be compared with the daily number of new COVID-19 cases identified by PCR testing. The ratio of these two values gives one daily values for the Infection Detection Efficiency (*IDE*), which can then be used to evaluate the performance of the testing program and the coupled contact tracing–testing program as means of monitoring and limiting the spread of the pandemic. One can also investigate which actions and/or COVID-19 pandemic events can be correlated with changes in the *IDE* value. The significant actions and events concerning the pandemic follow.

### 1.1 COVID-19 Pandemic History in Denmark into mid-2021

The first suspected COVID-19 patient in Denmark was positively diagnosed Feb. 26, 2020, with the first Danish COVID-19 hospitalization occurring Mar. 1, 2020. The 139 early Danish cases originated from travel to ski-resorts in Northern Italy and Austria [3]. Just as in many other European countries, travel-related cases were the initial sources of outbreaks that then spread rapidly in unprepared societies [4]. Denmark’s first wave of infection and hospitalization peaked at the end of Mar. into early Apr. 2020 [3]. Major non-pharmaceutical interventions, in particular lockdowns, brought the first wave of the pandemic under control, with cases being significantly reduced in Denmark and many other European countries [4]. Due to the success of the interventions, the imposed restrictions were loosened stepwise before the beginning of the summer holidays, late June 2020. However, in late Aug. and early Sept. 2020, a second wave of COVID-19 began, assumed to be caused mainly by young people returning from their summer holidays, as well as larger social gatherings. The same situation is described in many other European countries [4]. The pandemic in Denmark successively grew over the fall and the wave peaked in late Dec. 2020 (see Figs. 1, 2 in Section 2), declining rapidly thereafter due to a hard lockdown being reinstated Dec. 25, 2020. A new more virulent COVID-19 variant (B.1.1.7, the Alpha variant) began to spread in Denmark in Dec. 2020 and by Mar. 2021 was responsible for most of the COVID-19 infections in Denmark [5].

**Fig. 1:**
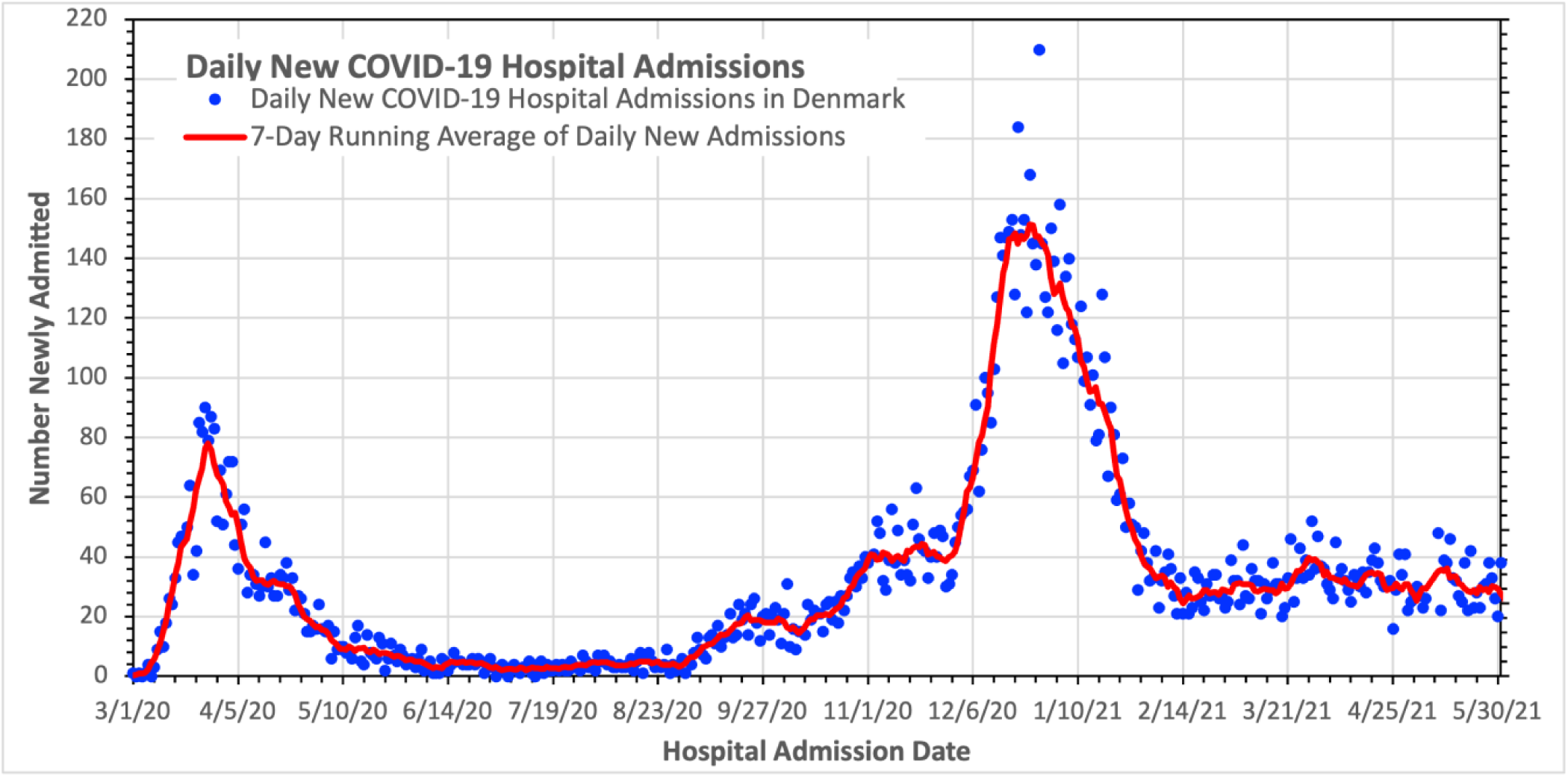
Number of daily new hospital admissions in Denmark. The points are actual values while the line traces out the 7-day running average values. Horizontal axis tick marks are at 1-week intervals.

**Fig. 2:**
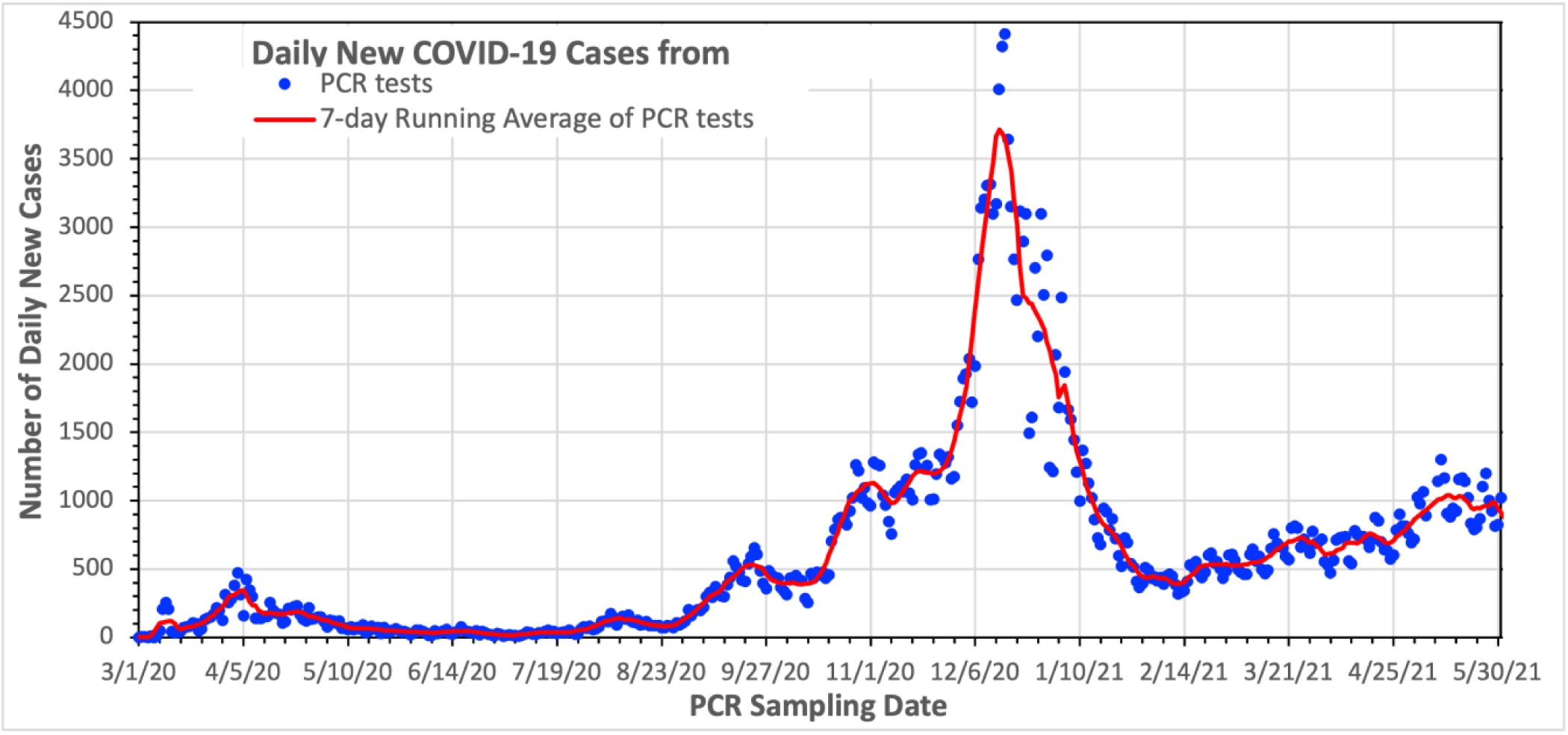
Daily number of new positive PCR tests in Denmark. “New” means that the individuals haven’t tested positive in the previous 60 days. The points are the actual values while the line shows the 7-day running average values. The date value is when the PCR sample was collected. Horizontal axis tick marks are at 1-week intervals.

Pharmaceutical intervention by vaccination began Dec. 27, 2020, the vaccinations starting with Denmark’s most vulnerable population, largely the elderly and healthcare workers. As of early Mar. 2021, some 8.3% of Denmark’s ∼5.83 million population had received at least one mRNA vaccination dose, while about 3.1% had received 2 doses as of that date. By early May 2021, ∼25% of the population having received at least 1 dose [6,7].

Below we summarize the main events of the pandemic in Denmark through early May 2021, which might be expected to show up in the calculated values of the *IDE* which are plotted in Section 3 and discussed in later parts of this paper. The full history of the major pandemic events in Denmark is chronicled in Ref. [8].

A hard lockdown was imposed in Denmark on Mar. 11, 2020, to bring the previously freely expanding pandemic under control. This lockdown was successively relaxed with a series of slow controlled re-openings in the late spring and early summer of 2020. Due to varying local conditions, localized COVID-19 outbreaks still occurred in some regions. Because of the low infection numbers in the country as a whole during the summer of 2020, these localized outbreaks were even seen in the national numbers. Fortunately, the localized outbreaks were relatively easily contained with local actions. Out of the five local outbreaks between Apr. 10 and Aug. 31, 2020, the largest was in Aarhus. It started in late July and lasted until the latter part of Aug. 2020. This outbreak was largely confined to the younger healthy population and as such there were relatively few cases that resulted in hospitalizations during this outbreak.

The re-opening of bars and the resumption of nightlife in mid-Aug. 2020 caused a major national infection increase that was somewhat damped by some re-instated restrictions in late Sept. 2020. However, colder weather and more indoor activities, together with significant travel activities due to the national fall holiday in Oct. 2020, caused a further infection increase. In addition, a regional outbreak in Northern Jutland occurred in late Oct. and early Nov. 2020. This outbreak was in part driven by the spread of COVID-19 among mink-farms and their employees with spillover of mink variants of COVID-19 to the surrounding society in North Jutland (around 4,000 people were estimated to have been infected). Northern Jutland was placed under a hard lockdown in early Nov. 2020. Furthermore, based on a Nov. 3, 2020 Statens Serum Institut (SSI) report [9], the Danish government decided that the entire mink population in Denmark (∼17 million) should be culled. This, together with enhanced and more focused testing along with the local lockdown helped to temporarily limit a further growth of the pandemic in the region. However, the stepwise growing pandemic in the remainder of the country over the fall of 2020 eventually resulted in a second major infection wave that peaked in Dec. 2020 and led to a second full national lockdown starting Dec. 25, 2020, which brought the infection wave under control. However, during Feb. and early Mar. 2021 Denmark again saw a steady increase in the infection level, in part due to the emergence of the more contagious B.1.1.7 (Alpha) variant (that was initially detected in Nov. 2020) and in part due to a slow reopening of society in late Feb. 2021 with the second lockdown ending Feb. 28. Thereafter restrictions continued to be eased. In the next subsection the testing and contact tracing effort in Denmark is reviewed in more detail.

### 1.2 Review of the Danish National COVID Testing/Tracing Program

Establishment of a Danish national testing effort, Test Center Denmark, was discussed in Mar. 2020, and started working in Apr. 2020. The SSI organized and published data of the day-to-day testing activities. Initially there were fewer than a thousand PCR tests/day, but this number was increased with time. By Aug. 1, 2020, the daily test capacity had reached 32,000 tests/day [10] distributed across the country (∼ 0.5% of the population per day). On Sept. 29, 2020, 50,817 PCR tests were given, of which 50,598 were taken by people who had not previously tested positive and with 4,512 testing positive for the first time. Also, by Sept. 29, 2020, a cumulative total of 3,841,072 tests had been given. By Dec. 14, 2020, testing levels reached 75,000 tests/day (∼ 1.3% of the population per day). In spite of this seemingly large size, there were reports (see e.g. [11,12]) of long wait times (multiple days) for testing in Dec. 2020, especially in the larger cities, when the pandemic was again spreading readily. In 2021, the daily number of PCR tests given continued to increase. As of Mar. 5, 2021, a total of 155,247 PCR tests were given on that day alone (∼ 2.7% of the population per day), with a cumulative total of 17,750,319 PCR tests having been performed in Denmark. This is approximately 3 tests on average for every individual in Denmark. Through Mar. 5, 2021, the accumulated number of PCR identified infected was 216,509 individuals (3.7% of the Danish population) [13]. Information about the number of PCR tests and Rapid Antigen tests performed and their results is available on the SSI Overvågningsdata website [13].

In addition to the PCR tests, Rapid Antigen (RA) tests were introduced and conducted by a few private companies (e.g., Falck, Carelink, and Copenhagen Medical). These tests were started in earnest in Dec. 2020. Through Mar. 5, 2021, 776,138 RA tests had been given, of which 5,184 yielded people who tested positive for the first time [13]. (It should be noted that any positive RA tests had to be confirmed via PCR testing in order to be counted as a COVID-19 case in the SSI database [13].) An examination of the global database of Ref. [14] shows that by the spring of 2021 Denmark had developed one of the world’s most ambitious testing programs in terms of the number of tests conducted per million individuals.

The Danish institution, Styrelsen for Patientsikkerhed (STPS), which is responsible for contact tracing, announced the beginning of a new more robust contact tracing program in June 2020 [15]. The program was successively expanded through the fall and winter of 2020/2021 to better deal with the growing number of COVID-19 cases. The data for the day-to-day contact tracing activities from the STPS reported herein were obtained through personal communication [16]. The contact tracing effort run by STPS seeks to identify the people that currently infected individuals had been in contact with for durations deemed “long enough” and proximity “close enough” to themselves potentially become infected (i.e. close contacts). It is assumed that all identified close contacts will be tested for COVID-19 and undergo isolation if they test positive. In addition to the STPS contact tracing effort there will be a spontaneous effect in which members of the general public that are infected will contact their acquaintances that they have been in contact with and suggest that they should get tested. The scale of this effect is unknown but it will supplement the STPS effort as it also reduces further transmission.

By early Dec. 2020, STPS had a taskforce of about 500 people conducting the national contact tracing, and by Dec. 17, 2020, this taskforce was expanded with about 800 additional personnel. The goal of this expansion was to be able to trace close contacts of 5,000 newly infected per day [17]. The scale of the operation growth is given by the following examples: on Sept. 29, 2020, STPS opened 520 cases for tracing their contacts, where 68 individuals out of the 520 infected could not be reached, and one infected individual did not want to participate. On this same day 1,100 close contacts were identified for follow up [16]. On Dec.14, 2020, STPS investigated 3,850 infection cases, where 564 could not be reached, while 10 did not want to participate, and 9,184 close contacts were identified for follow up [10]. On Mar. 5, 2021, STPS investigated 586 cases, where 17 could not be reached, and 4 did not want to participate. On that same day, 1,479 close contacts were identified for follow up [16]. For the time period ending in late spring 2021, the highest number of daily infected cases were opened by STPS was 5,532 and occurred on Dec. 19, 2020, while the highest number of close contacts identified for follow up was 12,185, which also occurred on Dec. 19, 2020 [16]. The average number of STPS identified close contacts per infected person was 2.6 [16]. The same Ref. also indicates that only about 29% of the *identified* infected in Denmark self-report no symptoms, while 66% self-report some symptoms, and 5% did not provide input. While this no-symptom-fraction is smaller than other values reported earlier (i.e., via prevalence surveys), it is biased in that it fails to account for asymptomatic cases that were never identified by contact tracing. In the next section the raw data used herein are presented.

## 2. Raw data used

The Statens Serum Institut (SSI) [18] maintains a large historic data set on the COVID-19 pandemic in Denmark [19]. From within the website [19], it is possible to download a variety of data files concerning the pandemic that cover a wide range of dates and topics. In particular, the files contain the history of the pandemic on a day-by-day basis up through the date of the chosen file. In exploring the data files, it was noted that values in older files were often updated in newer versions of the files to correct for mistakes, omissions, new procedures, etc. Thus, we chose a file that provided data from the beginning of the pandemic to well past the period of interest. To be specific, we arbitrarily chose *overvaagningsdata-covid19-01062022-qaoz.zip*, [13] corresponding to June 1, 2022. It is a compressed file that yields a folder [*overvaagningsdata-covid19-01062022-qaoz*] which has files that contain the data used in several of the subsections below.

### 2.1 Hospital admissions (Raw and 7-day running average)

The file “*Newly_admitted_over_time.csv*” from within the zip file [13] identified above contains the daily new hospital admission values. “Admissions” are stays of over 12 hours. The values from the “Total” column in the file are plotted in Fig. 1 as a function of time from the beginning of the pandemic to late May of 2021. As the values have large daily variations due to the relatively small size of a number of the values, the 7-day running average of the values is also plotted. A 7-day average was chosen in order to smooth out day-of-the-week effects.

### 2.2 New Positive PCR Tests (Raw and 7-day running average)

The file “*Test_pos_over_time.csv* from within the zip file [13] identified above contains the daily new positive PCR test number values. The values from the “NewPositive” column of the file are plotted in Fig. 2 as a function of time from Mar. 1, 2020 into late May of 2021. “New” means that they haven’t tested positive in the previous 60 days. Because of the large daily variations of the values within the file due to the relatively small size of a some of the values, as well as day of the week effects, the 7-day running averages of the values are also plotted to smooth out the day-of-the-week effects, as well as statistical error effects. The date value is the day when the PCR sample was collected.

### 2.3 COVID-19 Antibody Prevalence Values

In the time frame corresponding to the period of interest covered by this paper, multiple investigations of the prevalence of COVID-19 antibodies in the Danish populations were carried out. COVID-19 antibody prevalence values give the integral number of past COVID-19 cases in Denmark. This is because the COVID-19 anti-bodies are long lived enough (given the high sensitivity of the antibody tests used) to remain detectable throughout the period studied. Also, it takes about 14 days from infection for antibodies to become detectable [20–23].

Two series of studies of the COVID-19 antibody prevalence values in Denmark were done. The first was by SSI with results reported in [2,20–24]. The values reported therein were corrected for the sensitivity and a specificity of the Wantai SARS-CoV-2 Ab ELISA antibody test [25] used for the measurements. A few of the values were further corrected for gender, age, location, etc. to match the characteristics of the Danish population as a whole. The second series of studies was done by the Danish Blood Bank on Danish blood donors [26,27] and was also corrected for the sensitivity and a specificity of the used Wantai antibody test [25], but not for Danish population characteristics. The values from the 2 sets of studies are reported in Fig. 3 including their 95% confidence intervals. The data in the studies were given as percent values and were multiplied by the population values for Denmark interpolated between the 2 closest quarterly values from Ref. [7] for the quoted date of the antibody sample collection. (The population values used range from 5,824,857 for Apr. 1, 2020 to 5,850,189 for July 1, 2021. [7]) The blood donor values are typically weekly in nature for which the corresponding mid-day of the Danish week were used as the sampling date (i.e., Thursdays), while the SSI values were either weekly or several weeks in duration with the median date of the sample collection being used as the date for the plotted results.

**Fig. 3:**
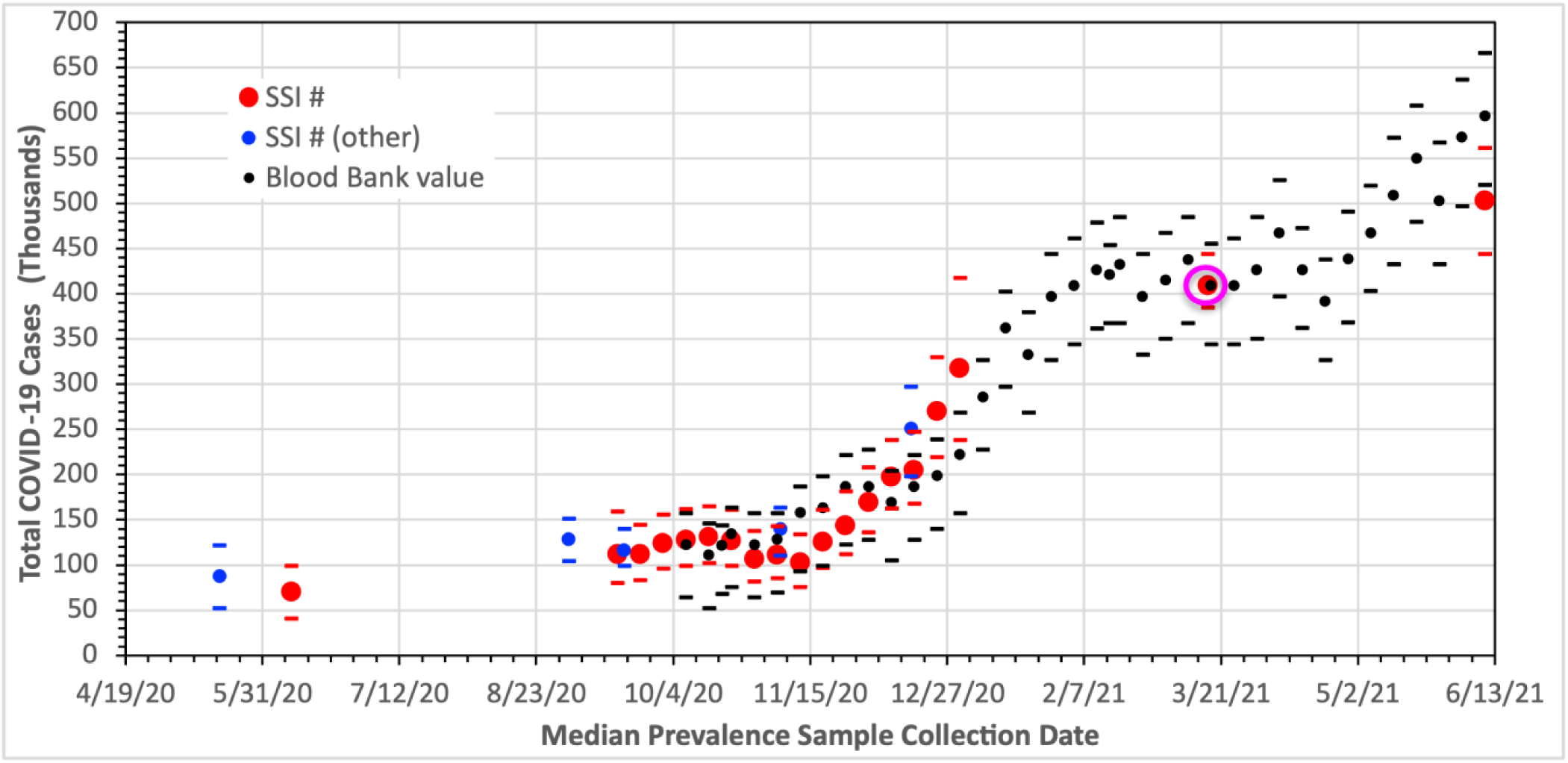
Measured Danish antibody prevalence results over time. The dots are the values while the bars represent the associated lower and upper 95% confidence intervals. The prevalence values were determined using the Wantai ELISA antibody test [25] and all values were corrected for the test’s sensitivity (0.97) and specificity (0.995). The red and blue dots are SSI results [2,20–24], the blue dots being further corrected for participation biases. The black dots are data [26,27] from Blood Bank donors. The data are given by the Refs. as percent values which we multiplied by the Danish population values to get the number of cases as noted in the text. The time axis gives the assigned dates of the prevalence values as discussed in the text. Horizontal axis tick marks are at 1-week intervals. The Round 4 value indicated by the red dot in the magenta oval is the value used to do the scaling in Section 3.3.

### 2.4 Alpha variant

The characteristics of the pandemic changed with time as different variants of COVID-19 emerged. During the period covered by this paper, the only “one” that had a significant impact on the results presented here is the Alpha variant. It first appeared in Denmark in Nov. of 2020 and quickly became the dominant strain during the first half of 2021. The Alpha variant is more virulent. The impact and adjustments due to the Alpha variant on our results are given in Section 3. The percentage of COVID-19 cases that are due to the “Alpha” variant are shown in Fig. 4, the values being from Refs. [5] and [28]. Before Nov. 12, 2020, an “Alpha” % value of 0 is used.

**Fig. 4:**
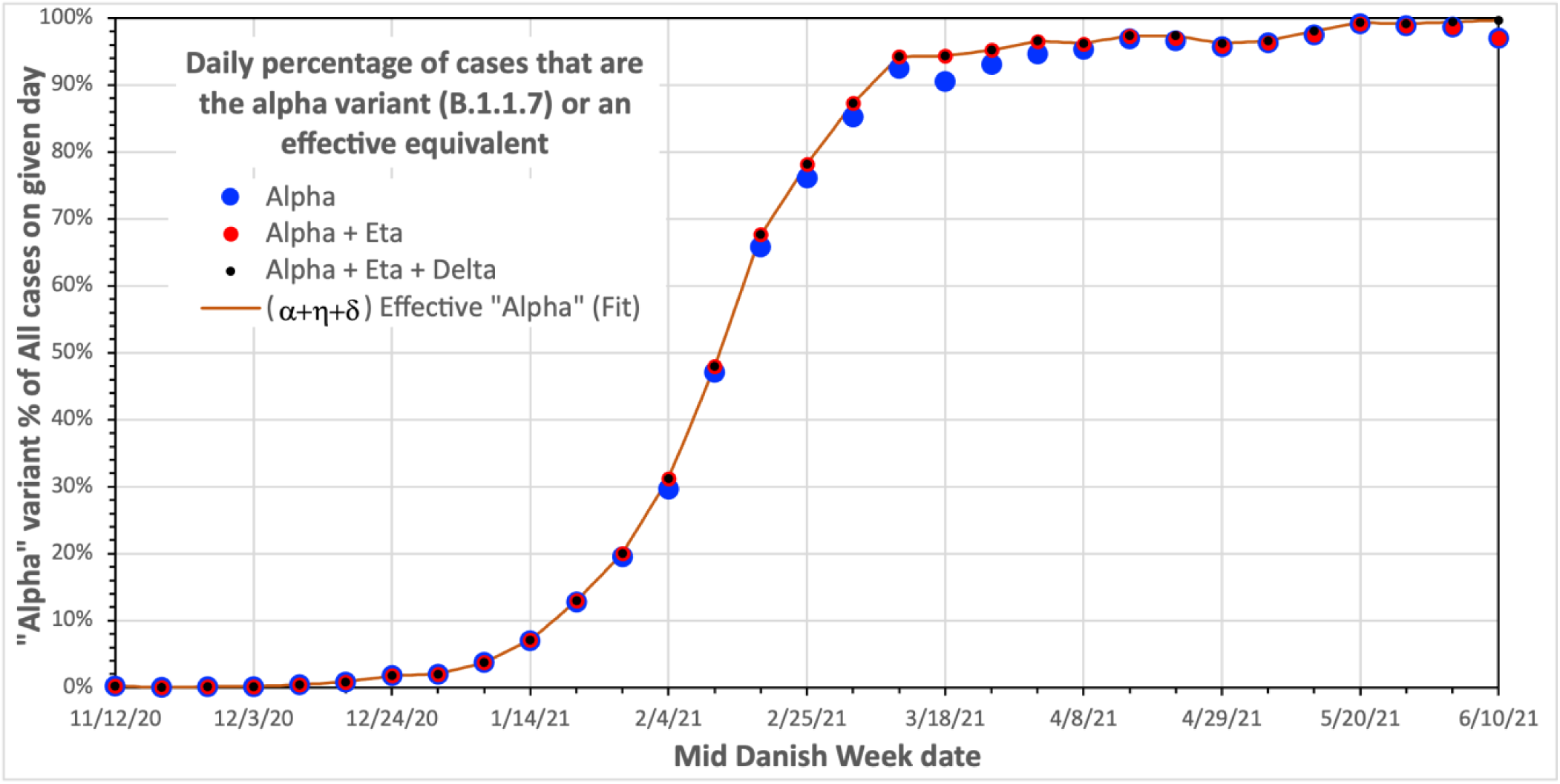
Measured fraction of Danish COVID-19 cases due to the more virulent “Alpha variant” [5,28]. As seen in the legend, we include the Eta variant which peaks at 3.8% on Mar. 18, 2021 and the Delta variant whose maximum value in the plot is 2.8%, which occurs on June 10, 2021 as part of the “Alpha variant”. The values are given by week number in the above references, which we assign a value of the Thursday in the corresponding week number (i.e., the Danish week running Monday through Sunday). The curve is the linear interpolation between the “Alpha *+ Eta + Delta %” points. The Fit is used in Section 3. Horizontal axis tick marks are at 1-week intervals*.

In reality, as noted in Fig. 4, two other variants appeared in the time period covered. These were the Eta and Delta variants. Both are relatively minor contributors in the time frame shown; the Eta variant peaked at 3.8% of all new infections on Mar. 18, 2021 while the Delta variant had a maximum value in the plot of 2.8% which occurred on June 10, 2021. As an aside, it is noted that in the period beyond that shown in the plot, the Delta fraction continues to grow, becoming the dominant variant in July 2021. The Delta variant is somewhat more virulent than the Alpha variant [29]. We were unable to find firm data on the relative virulence on the Eta variant. However, even though it never became a major fraction, it did grow in percentage values during the period where the Alpha variant was rapidly expanding. Thus, it was assigned a disease seriousness equivalent to that of the Alpha variant.

## 3. Infection Detection Efficiency

We define the Infection Detection Efficiency (*IDE*) on day *t* as the following ratio:

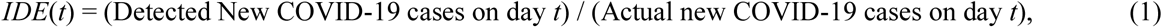

where (New COVID-19 cases detected on day *t*) is the number of new cases of COVID-19 cases that are detected and/or verified via PCR tests. The denominator is self-explanatory, but as pointed out in the Introduction, its value has to be determined from a stand-in as its value on a daily basis is not in practice measurable. A scaled version of new daily hospital admissions is used as the stand-in, the scaling being done using the COVID-19 antibody prevalence values. In calculating a value for *IDE*(*t*) several issues must be addressed. These are discussed below.

### 3.1 Timing Issues

In calculating the *IDE*, one has to ensure that the values used for the numerator and denominator of Eq. (1) are properly matched in time. To do this we need to have values for the different stages associated with a COVID-19 infection. For an infection, we assume that the initial exposure occurs at time τ = 0. There is a considerable variation on when symptoms appear in COVID-19 infections (if they appear at all) and published mean times of symptom appearance differ as noted in review articles (e.g. Refs. [29,30]). Furthermore, according to Ref. [31], the time from infection to the appearance of symptoms became shorter for new COVID variants. For the original “wild-type” strain, Ref. [29] reports a mean symptom appearance time τ_symptom_ = 6.58 days, with Ref. [30] reporting a mean {median} value of τ_symptom_ = 5.8 {5.1} days for the original strain. For the newer strains Ref. [31] reports mean {median} values of τ_symptom_ = 5.0 {5} days (Alpha), τ_symptom_ = 5.1 {5.0} days (Beta / Gamma), and τ_symptom_ = 4.3 {4.0} days (Delta). Of the newer strains, the only one having a significant contribution to the <u>integral</u> number of COVID-19 cases into early June 2021 was the Alpha variant and through that time it only made up about 30% of the <u>integral</u> number of cases. Also as of early Mar. 2021, the <u>integral</u> Alpha contribution was only about 6% of the total number of cases. (Note that Fig. 4 gives daily values and not the integral values.)

Considering the above set of values and the fact that most of the measured values we use in calculating the *IDE* are for whole days, we decided to use a value of τ_symptom_ = 6 days regardless of date (or variant). We also assumed that people will come in for PCR testing on the same day as symptoms appear and therefore, we assigned the same time interval from exposure to PCR testing, namely τ_PCR_ = 6 days.

Next, we consider the delays from viral exposure to the development of measurable values of antibodies (positive results) for the prevalence tests. The SSI prevalence reports/papers [2,20–24] state that it takes approximately 14 days from exposure to the virus until measurable antibody levels appear. Herein we adopt their value, namely τ_prevalence_ = 14 days.

Finally, we examine how many days elapse from virus exposure until hospital admission for those individuals that are hospitalized due to COVID-19, namely τ_hospital_. The literature either directly reports values for τ_hospital_ or instead the times from symptom onset to hospitalization, in which case one has to add τ_symptom_ = 6 days to the symptom-onset based hospitalization values. There is a considerable variation in the reported τ_hospital_ values from different sources. Part of this is due to differences in what is deemed to be hospital admission. In Denmark hospital admission means hospital stays that are longer than 12 hours and the admissions are for clinical reasons and not for isolation purposes. In contrast, papers from the UK, Singapore, and China often refer to hospitalization as any COVID-19 related contact with hospitals or admission for isolation purposes as opposed to for clinical need. As such, these latter source values are ignored.

A large investigation in terms of cases studied (14,618 hospitalized COVID-19 patients) was done in Belgium [32] and reported a median value of 5.0 days from symptom onset to hospitalization, yielding a median value of τ_hospital_ = 11 days. This is the value that will be adopted in this paper. (A much smaller study in northeastern Germany involving 36 hospitalized patients who provide information on symptom history yielded a median value of 6 days from symptom onset to hospitalization [33].)

The above set of values, i.e., τ_symptom_ = τ_PCR_ = 6 days, τ_hospital_ = 11 days, and τ_prevalence_ = 14 days, imply the following:

- When comparing the integral number of antibody prevalence cases with the integral number of hospital admissions, one has to use a value 3 days earlier (*i.e.* τ_prevalence_ – τ_hospital_) for the hospital admission integral than the date of the “corresponding” prevalence value.
- When comparing the daily number of new COVID-19 cases from the PCR data with the daily number of new COVID-19 cases as determined from the scaled daily new hospitalization values, one has to use a date 5 days later (i.e. τ_hospital_ – τ_PCR_) for the hospital admission value than for the date of the “corresponding” PCR value.
- When comparing other values one may need other shifts.
- When results are plotted, one must note which quantity the date axis refers to.

### 3.2 Alpha variant adjustments

The ability to use a quantity such as the daily number of new hospital admissions as a stand-in for the actual number of COVID-19 cases in Denmark requires the use of a known scaling factor between the two quantities. Also, due to the shifts identified above, if we use the time of PCR testing as a reference, then we have (actual new daily infected (*t*_PCR_)) = (daily new hospital admissions (*t*_PCR_+5)) × (scaling factor) due to the 5-day delay between the average time of PCR testing and the time of hospitalization. Ideally, the scaling factor would remain constant in time. Unfortunately, this is not the case. According to a study in Denmark [5], on a per COVID-19 case basis, the Alpha variant results in a factor of 1.42 (1.25–1.60, 95% CI) greater likelihood for hospitalization than the initial “wild-type” non-Alpha COVID-19 versions after correction for factors such as age, sex, Denmark regions, comorbidity, etc. Other studies {e.g. [34,35]} show similar results of higher hospitalization for Alpha variant cases, 1.71 (1.64–1.79, 95% CI) and 1.52 (1.47–1.57, CI) in Sweden and the U.K. respectively. For the work herein, as it deals with COVID-19 in Denmark, we will use the value of 1.42 from Ref. [5], which is also a Danish result. We will also assume that the other variants that follow the Alpha variant in time (Eta and Delta) have the same adjustment factor as the Alpha variant. While not totally true, the error introduced is negligible given the small contribution that the Eta and Delta variants make relative to that of the Alpha variant in terms of cases in the time period of interest covered herein. Thus, we now have: (actual new daily infected (*t*_PCR_)) = (daily new hospital admissions (*t*_PCR_+5)) × (a time varying scaling factor (t*ime*)), where *time* is the time with the appropriate shift.

To “correct” for the higher probability of hospital admission for Alpha variant cases, the time varying part of the scaling factor can be accounted for by appropriately adjusting the hospital admission numbers. Thereby we can use a constant scaling factor to yield values for the estimated daily number of new COVID-19 cases using the values shown in Figs. 1, 3, and 4. The constant scaling factor chosen is the one corresponding to the “wild-type” COVID-19 hospitalization admissions; namely the hospitalization admission values involving “Alpha variant” cases were adjusted to correspond to what would have been expected if only “wild-type” cases were involved.

Let *H*(*t_hospital_*) be the number of reported new hospital admissions on date *t_hospital_*, the values being shown in Fig. 1. Let *HA_Alpha variant adjusted_* (*t_PCR_*) be the “Alpha variant” adjusted number of new hospital admissions that are also appropriately time shifted to directly correspond to the PCR sampling date. Using adjusted hospital admission values allows the use of a constant scaling factor to convert adjusted hospital admission values to actual COVID case numbers for all times and also has the needed time shift built in. Let *F_α_* (*t_PCR_*) be the fraction of the COVID-19 cases that are due to the “Effective Alpha” variant on date *t_PCR_*, namely the fractional values corresponding to the solid curve in Fig. 4. (It is assumed that the collection time of samples taken for variant determination is the same as for PCR sampling.) We take *F_α_* (*t_PCR_*) = 0 prior to Nov. 12, 2020 and *F_α_* (*t_PCR_*) = 1 after June 10, 2021. Finally, let *C* (= 1.42) [5] be the fractional increase in hospitalization for the “effective Alpha” variant cases relative to that for the “original wild-type” cases. We then have

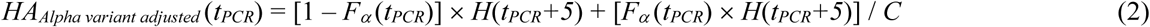

where the first right hand side term in the sum is the contribution of the “original wild-type” cases to the adjusted hospitalization values and the second term is that for the “Effective Alpha” cases for date (*t_PCR_*). Explicit use of the assumption that the hospitalization times and PCR times differ by 5 days (i.e. *t_hospital_* = *t_PCR_* + 5) is made. The daily values and 7-day running averages of *HA_Alpha variant adjusted_* are plotted in Fig. 5 as they are for *H* for comparison.

**Fig. 5:**
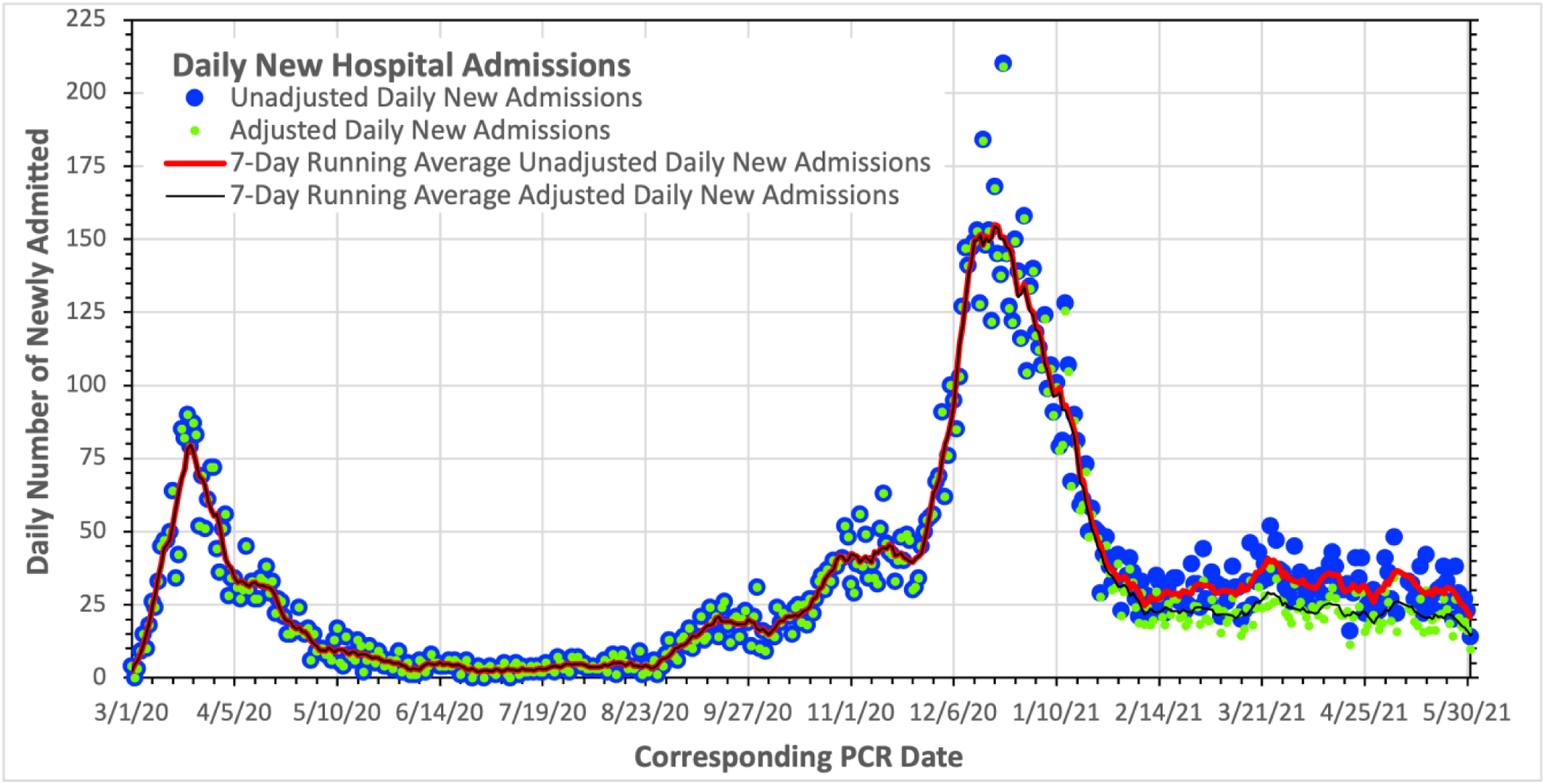
The number of daily new hospital admissions due to COVID-19 in Denmark. The points are the daily values while the solid curves give the 7-day running average values. The Unadjusted and the Adjusted values are shown. The adjustments allow for the use of a single constant scaling factor throughout the time frame shown to convert COVID-19 hospitalization values to actual COVID cases in Denmark. The adjustments account for the fact that the “Effective Alpha” variant (“Alpha + Eta + Delta %”) results in fractionally higher hospitalization per COVID case than the original COVID-19 strains. As expected, this means lower “Adjusted” values relative to the Unadjusted values in the region where the “Effective Alpha” related variants play a noticeable role (starting in 2021). Horizontal axis tick marks are at 1-week intervals. Relative to Fig. 1, the curves and points are shifted to the left by 5 days as the dates used are that of when the corresponding PCR samples for the hospital admissions are assumed to have been collected (i.e., 5 days before the admission dates). Also note that the Unadjusted values are the same as the values gotten by setting F_α_ (t_PCR_) = 0 for all times in Eq. (2).

### 3.3 Scaling Factor

Using the values for *HA_Alpha variant adjusted_* (*t*) from the previous section and antibody prevalence values from Section 2.3, a value for the scaling factor needed to convert the adjusted daily new hospitalization numbers into daily new COVID cases is calculated. To do this properly, one must consider the timing issues discussed in Section 3.1 and use the prevalence results. As the prevalence values reflect the integral number of COVID-19 cases, we chose a value from late within our period of interest, but before the impact of vaccinations became significant. A late choice yields a value more representative of the entire period being studied and should also yield a larger number and fraction of positive cases and thus better-quality results. Furthermore, an SSI value was chosen as opposed to Blood Bank values due to higher statistics for the SSI studies and because the SSI studies were designed to be representative of the Danish population as a whole, which was not true of the Blood Bank values. (It should however be noted that as seen in Fig. 3, the SSI and Bloodbank values are in agreement within their error bars.)

In particular, we chose the Round 4 prevalence results [7.0%, with a 95% confidence interval (CI): 6.6-7.4%] which had a median sampling date of Mar. 17, 2021 [21]. As per Section 3.1, this value needs to be compared with the integral of the daily new hospital admissions through a date 3 days earlier, namely Mar. 14, 2021. Also, as per Section 3.2, the integral should use the corresponding “Adjusted” daily new hospital admission values *HA_Alpha variant adjusted_* (*t_PCR_*), through *t_PCR_* = Mar. 9, 2021 given that PCR values correspond to admissions 5 days later. That integral has a value of 12,758.

The prevalence numbers are given as a percentage of the Danish population. To convert these values into case numbers we multiply the percent value/100 by the interpolated population values between the 2 closest quarterly values from Ref. [7] for the related date, just as in Section 2.3. This yields 408,996 COVID-19 cases for the Mar. 17, 2021 prevalence value. Taking the ratio of this prevalence value to the integral of the “Adjusted” new hospital admission values from the preceding paragraph yields a scaling factor *f_scale_* of 408,996/12,758 = 32.058 being needed to convert the daily “Alpha variant” adjusted new hospital admissions into the daily number of new actual COVID-19 cases. Thus, we have (daily new COVID cases (*t_PCR_*)) = *f_scale_* × (*HA_Alpha variant adjusted_* (*t_PCR_*)). As the 95% CI for the prevalence value of 7.0% spans the range ± 0.4/7.0 or a percentage interval of ± 5.71% of the central value, the 95% confidence interval of the scaling factor would have the same percentage interval, or *f_scale_* = 32.058 ± 1.832.

As an aside, it is noted that the use of the reported versus the “Adjusted” integral hospitalization values would cause only a minor change in the scaling factor. This is because in our main time period of interest through early May 2021, the Alpha variant contributes to the case count only when there are relatively few COVID-19 cases. More specifically, the reported integral hospitalization count corresponding to the Mar. 17 prevalence date is 13,033 versus 12,758 for the “Adjusted” integral hospitalization count. This would have resulted in an approximate two percent lowering of the scaling factor value.

### 3.4 Infection Detection Efficiency Results

With a value for the scaling factor in hand, it is now possible to convert the daily “Adjusted” new COVID-19 hospital admission values into daily new COVID-19 cases values as (daily COVID cases (*t_PCR_*)) = 32.058 × (*HA_Alpha variant adjusted_* (*t_PCR_*)). In Fig. 6 these values and their 7-day running averages are plotted. Also plotted in Fig. 6 are the daily new PCR positive values shown in Fig. 2 along with their 7-day running average values.

**Fig. 6:**
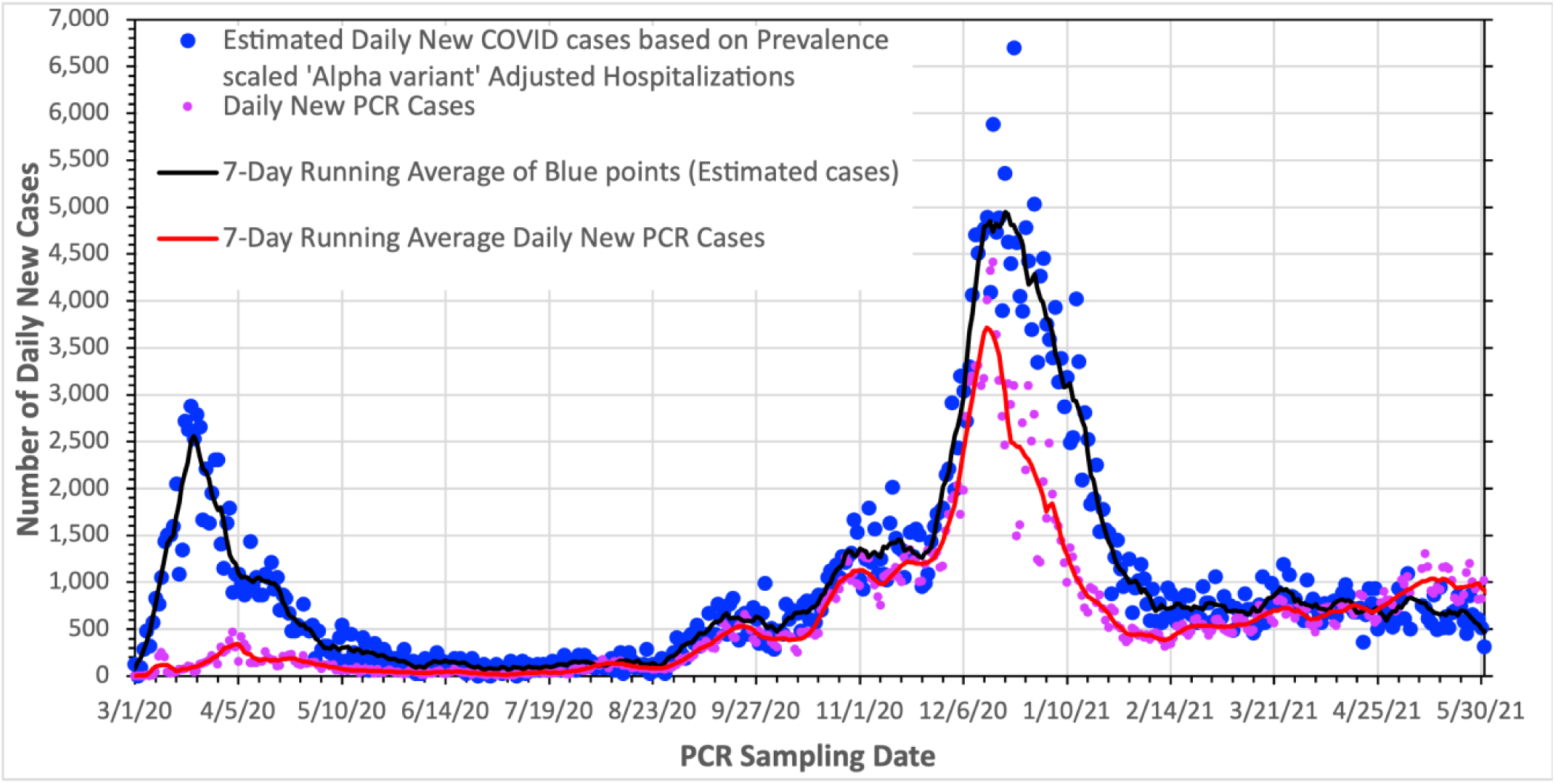
The number of daily new COVID-19 cases in Denmark as given by the Prevalence scaled “Alpha variant” adjusted new hospitalization values and the PCR measurements. The points are the daily values while the lines represent the 7-day running average values. The date values correspond to when the PCR samples were taken. The hospitalization derived values make use of HA_Alpha variant adjusted_ (t_PCR_), and thus already have the needed 5-day shift built in. The daily New PCR case values are also shown. Horizontal axis tick marks are at 1-week intervals.

From the values shown in Fig. 6, the values of *IDE* can be calculated using Eq. (1), expressed as (reported COVID cases (*t_PCR_*)) / (daily COVID cases (*t_PCR_*)). However, before plotting the actual *IDE* values, it is immediately clear from examining the curves in Fig. 6 that there will be issues. First, through the spring of 2020 the testing capability in Denmark is completely inadequate to deal with the scale of the pandemic and there is no point in even examining the *IDE* values until the summer of 2020. Second, starting in mid-Apr. 2021, the number of PCR detected cases is greater than the actual number of COVID-19 cases, a physical impossibility. A close look at Fig. 6 also shows that this same problem occurs for a few days early in Aug. 2020. Finally, starting late in 2020 and extending in Feb. 2021 the ratio of the curves seems unusually low. The plot of the *IDE* values in Fig. 7 clearly shows these issues.

**Fig. 7:**
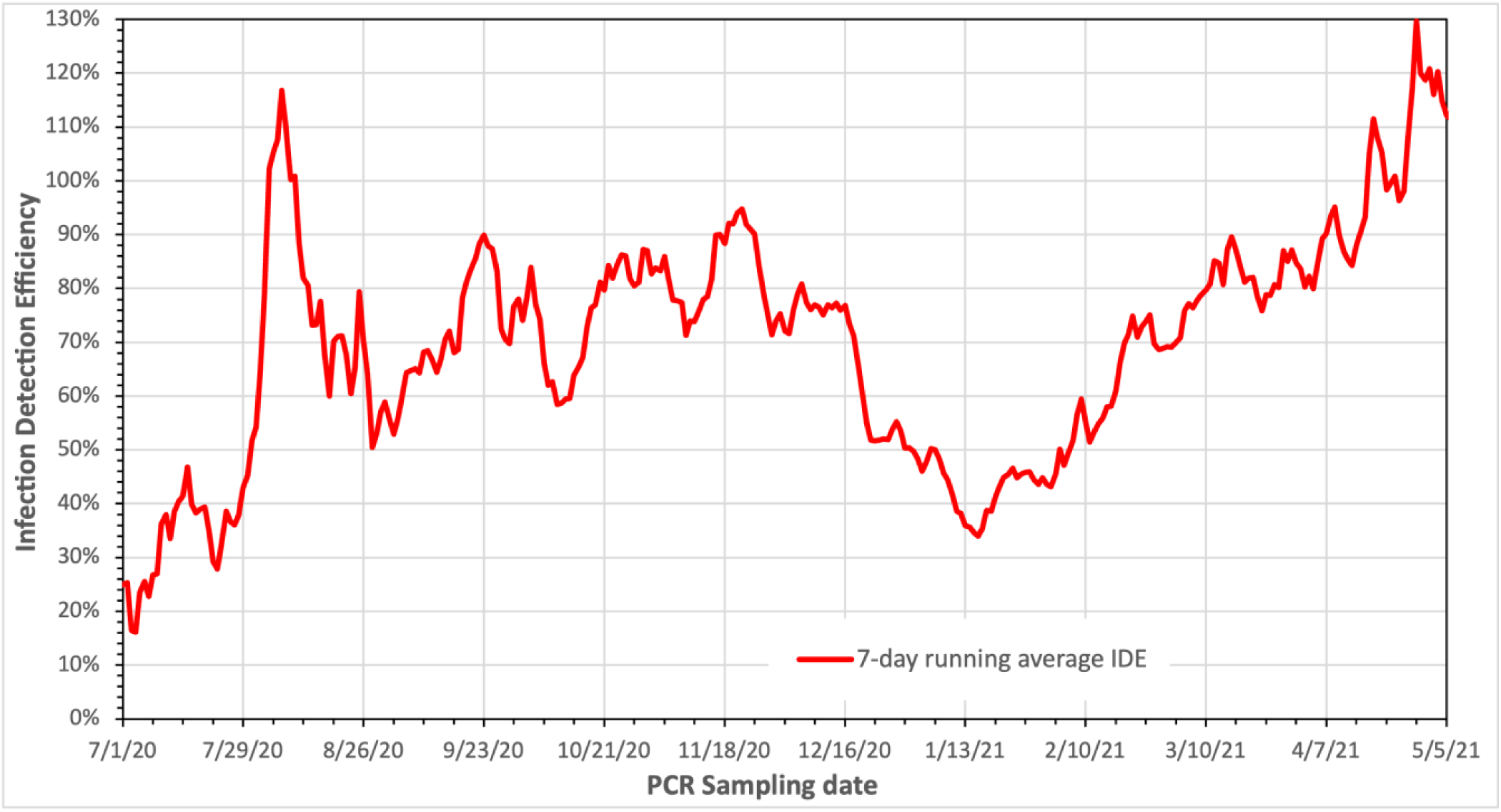
The 7-day “running average” daily values of the Infection Detection Efficiency [Eq. (1)], but with its denominator calculated using the “Effective Alpha” variant corrected version of the hospitalization values given by Eq. (2) that are then scaled using the prevalence results. The unphysical (>100%) values are discussed in the text. The “running average” values are the 7-day running average of the numerator of Eq. (1) divided by the 7-day running average of the denominator of Eq. (1). Horizontal axis tick marks are at 1-week intervals.

Concerning the unphysical values of the *IDE* at late times in Fig. 7, this can be attributed to the fact that by April and May 2021 a significant fraction of the elderly Danish population had been vaccinated against COVID-19, while only a small fraction of the younger population had received vaccinations. Prior to the COVID-19 vaccinations, the elderly comprised the greatest fraction of COVID-19 hospitalizations [36], but not of the number of COVID-19 cases. Thus, in April and May of 2021 the number of hospitalizations will be significantly reduced resulting in a corresponding reduction in the number of calculated COVID-19 cases. At the same time, the number of PCR cases are only marginally reduced as the elderly make up a much smaller part of the Danish population. The result will be faulty (> 100%) *IDE* values in this time frame.

In order to explore Fig. 7 and its associated issues in more detail, one will need to recall the details of the COVID-19 testing and tracking program development and performance (which the *IDE* is meant to assess) that were discussed in Section 1.2.

### 3.5 *IDE* Issues

As noted earlier, there are several issues associated with the *IDE* values shown in Fig. 7. The low but rising values of the *IDE* in the July 2020 and continuing into Sept. 2020 (with the exception of the large peak in Aug. 2020) are likely indicative of startup issues of the new contact tracing program. If one ignores the Aug. 2020 peak, it appears that the testing and contact tracing program does not achieve maturity until after the first week of Sept. 2020; the maturity being needed to achieve semi-stable *IDE* values. Furthermore, the unphysical values of the *IDE* (> 100%) at late times in the plot, were already attributed to age effects associated with vaccinations. The particulars of the other time period where the *IDE* values are > 100% (namely early Aug. 2020) are now examined in detail. That *IDE* peak can be correlated with a large, highly localized outbreak in the municipality of Aarhus at a time when there were very few cases in the remainder of the country. In the first half of Aug. 2020, the new COVID-19 cases in Aarhus accounted for up to 45 % of the new cases in the entire country (see Fig. 8) according to PCR testing results [13]. In contrast, the Aarhus population comprises only ∼ 6 % of Denmark’s population. Furthermore, the outbreak was largely confined to the younger population. This can be confirmed by examining the age distribution of PCR verified COVID-19 cases in Denmark during the time interval of the Aarhus outbreak and comparing it with the age distribution of COVID-19 cases in Denmark for identical time intervals before and after the Aarhus outbreak. The age distribution of the integral number of PCR identified cases is available from Ref. [37]. More specifically, files were downloaded in 3 pairs: July 1, 2020 & July 22, 2020; July 29, 2020 & Aug. 19, 2020; and Sept. 23, 2020 & Oct. 14, 2020. Choosing the “Cases_by_age.csv” files therein and subtracting the earlier date values from the later date values for each of those pairs yields the age sorted COVID-19 case numbers for each of the three 3-week intervals. These values are given in Table 1.

**Fig. 8:**
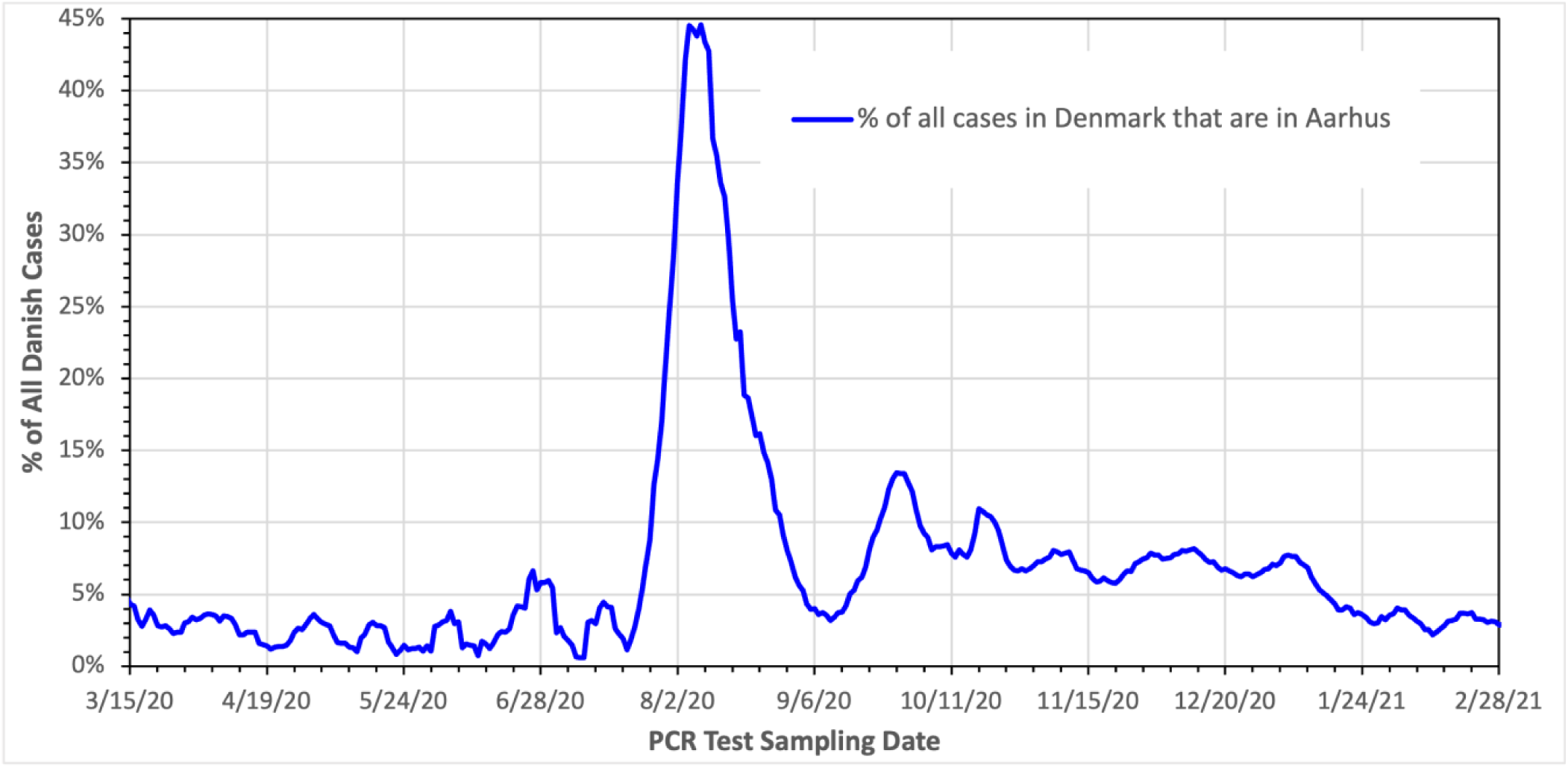
Daily values of the fraction of COVID-19 cases in Denmark that occur in the municipality of Aarhus which has 6% of Denmark’s population. The case numbers were determined by PCR tests. Horizontal axis tick marks are at 1-week intervals.

**Table 1.** The number of new Danish COVID-19 cases determined from PCR results per 10-year age group for 3 different 3-week periods. The middle period (grey) corresponds to time of the Aarhus peak and is strongly influenced by that peak as seen in Fig. 8. The two other periods [earlier and later (white columns)] are for comparison purposes. The ratios of the number of young patient cases (ages 0-39) to elderly patient cases (ages ≥ 60) are also calculated and presented. Ref. [37] gives integral values and the first row below gives the date values subtracted.

| Period | July 22 – July 1, 2020 |  | Aug. 19 – July 29, 2020 |  | Oct. 14 – Sept. 23, 2020 |  |
| --- | --- | --- | --- | --- | --- | --- |
| Age Group | PCR positive | % of positive | PCR positive | % of positive | PCR positive | % of positive |
| 0-9 | 29 | 5.216% | 138 | 5.984% | 535 | 5.793% |
| 10-19 | 73 | 13.129% | 391 | 16.956% | 1,693 | 18.330% |
| 20-29 | 115 | 20.683% | 545 | 23.634% | 1,955 | 21.167% |
| 30-39 | 81 | 14.568% | 376 | 16.305% | 1,155 | 12.505% |
| 40-49 | 91 | 16.367% | 316 | 13.703% | 1,356 | 14.682% |
| 50-59 | 79 | 14.209% | 325 | 14.094% | 1,283 | 13.891% |
| 60-69 | 42 | 7.554% | 130 | 5.637% | 649 | 7.027% |
| 70-79 | 27 | 4.856% | 65 | 2.819% | 414 | 4.482% |
| 80-89 | 10 | 1.799% | 16 | 0.694% | 158 | 1.711% |
| $\geq 90$ | 9 | 1.619% | 4 | 0.173% | 38 | 0.411% |
| Total of above | 556 | 100.000% | 2,306 | 100.000% | 9,236 | 100.000% |
| Old ($\geq 60$) | 88 | 15.827% | 215 | 9.324% | 1,259 | 13.631% |
| Young (0-39) | 298 | 53.597% | 1,450 | 62.879% | 5,338 | 57.796% |
| Young/Old | 3.386 | 3.386 | 6.744 | 6.744 | 4.240 | 4.240 |

In comparison with other periods, the Aarhus peak interval is different in the sense that it is predominantly confined to the younger part of the population. As can be seen in Table 1, the ratio (last row) of the number of the cases in the younger population (0 – 39 years old) to that in the elderly population (≥ 60 years old) is about twice as large during the Aarhus peak compared to its value in similar 3-week intervals before and after the Aarhus peak.

In general, the elderly make up a disproportionally large fraction of the hospitalized COVID-19 patients in Denmark, accounting for 69% of them through mid-May 2020 [36], but only account for ∼26% of Denmark’s population. As such, there will be a significant undercount of COVID-19 cases in Denmark during the period of the Aarhus peak as the number of cases is derived from the number of COVID hospitalizations, which is uncharacteristically low during this time period. This in turn results in unreasonably large *IDE* values given that the actual number of COVID-19 cases is the denominator of Eq. (1). This is just what is seen during the Aarhus peak (July 30 through Aug. 19, 2020 inclusive; namely the integral values of Aug. 19 minus those of July 29, 2020).

Another “strange” area of *IDE* values seen in Fig. 7 is for the period from mid-Dec. 2020 to mid-Feb. 2021 during which the *IDE* value drops to about 50%. In this interval there are several factors that might contribute to this. Early in this period, there was a very high demand for PCR testing which challenged the testing capacity at that time [11,12]. The demand was driven by requirements for testing prior to traveling (travel being driven by the upcoming Christmas and New Year holidays). This was aggravated by the large COVID-19 outbreak at that time as seen in Fig. 6. The high demand for testing and long lines likely resulted in frustration and people giving up on getting tested which likely led to some undercounting of COVID-19 cases by PCR and thus lower *IDE* values. However, the scale of this effect nor whether the above explanation is valid is not known. The large outbreak at that time also led to another nationwide shutdown starting Dec. 25, 2020 which continued through the end of Feb. 2021. The shutdown greatly reduced the infection scale by early Jan. 2021 and with the holidays over, the testing capacity was again more than adequate. Yet the low values of the *IDE* continued well into Feb. 2021 indicating that another or additional explanation is likely needed for the mid-Dec. 2020 to mid-Feb. 2021 low *IDE* values.

### 3.6 The Need for AGE-Based Adjustments

Having already noted the need to apply adjustments to the hospitalization values due to the following: the Alpha variant, age-related effects on hospitalization values causing unphysical *IDE* values for the Aarhus outbreak, and both vaccination and age-related effects also causing unphysical *IDE* values in the April – May 2021 time frame, it was decided to explore the age impacts more deeply. This was further driven by the strong correlation between age and the hospitalization likelihood that was previously noted.

In the example given below, the need for AGE-based adjustments is examined. The example examines 3 different scenarios which could also be viewed as values on 3 different days. In the example, the cases are subdivided into two groups: an Old age group and a Young age group. For each scenario, there are assumed to be 100 Known COVID-19 cases in total. (There could be additional Unknown cases that would be symptom free and thus don’t undergo PCR testing. Such cases would only be inferred/caught by prevalence results.) The difference in the 3 scenarios is in the number (or fraction) of Known cases that are in the Old age group versus the Young age group.

For the Normal or Average scenario, the Known cases are assumed to be equally divided between the two age groups, while for the other two scenarios the distribution is not equal. Furthermore, it is assumed that for the Old age group 80% of the cases that test PCR positive end up being hospitalized, while for the Young age group only 20% of those that test PCR positive end up being hospitalized. (The hospitalization fractions for each age group are assumed to remain constant over time or scenario.)

The situation and results for the 3 scenarios are given in the Table 2. As can be seen, although each scenario has the same number of Known COVID-19 cases, each scenario yields a different number of hospitalizations. Thus, unless the AGE-based effects can be adjusted for, the number of hospitalizations cannot be used to give the actual number of COVID cases so that accurate values for the *IDE* can be derived.

**Table 2.** The properties and results associated with the 3 scenarios in our example.

| Scenario | Known cases | PCR Young group cases; (fraction) | PCR Old group cases; (fraction) | Young group hospitalized (20% of cases) | Old group hospitalized (80% of cases) | Total hospitalized |
| --- | --- | --- | --- | --- | --- | --- |
| Normal or Average | 100 | 50; (0.50) | 50; (0.50) | $50 \times 20\% = 10$ | $50 \times 80\% = 40$ | 50 |
| Young group heavy | 100 | 75; (0.75) | 25; (0.25) | $75 \times 20\% = 15$ | $25 \times 80\% = 20$ | 35 |
| Old group heavy | 100 | 25; (0.25) | 75; (0.75) | $25 \times 20\% = 5$ | $75 \times 80\% = 60$ | 65 |

### 3.7 A Simple AGE-Based Adjustment

With known issues existing for the “Alpha variant” only adjusted *IDE* values and indications that age related effects could play a role, a Simple AGE-based adjustment was tried. Just as for the example in the preceding subsection, the COVID-19 cases were broken down into 2 age groups. The Old group was chosen to consist of all those of age ≥ 60 years old and by default the other group was those younger than 60 years old. This grouping was chosen since as noted in Section 3.5, those ≥ 60 years of age make up nearly 70% of all COVID-19 related hospitalizations, while at the same time are a relatively small fraction of the Danish population (just over 25%).

The AGE-based adjustments to the daily new hospital admission values require the use of daily values of the age distribution of new PCR verified COVID-19 cases. These values are available from Refs. [38,39]. The values from Ref. [39] are claimed to be sourced from the SSI who published the Ref. [38] values; the values being identical where they overlap. There are however several issues with the data values retrieved: **I)** There are gaps in the values that are typically associated with weekends and holidays. Somewhat surprisingly, there are fewer gaps for the Ref. [39] values. For the missing days, the gaps are filled in with linearly interpolated values utilizing values from immediately before and after the gaps. **II)** The values are integral numbers from the start of the pandemic rather than daily values. As such, values from successive dates were subtracted to yield daily values. **III)** Ref. [39] notes that changes were made in the collection/evaluation of the values for the dates of Mar. 10 and Mar. 25, 2021. An examination of the numeric values in and around this time frame shows a sudden jump in the integral values on Mar. 10, 2021 followed by a sudden drop on Mar. 25, 2021 back to values more in line with the trend seen prior to Mar. 10, 2021. Given the inconsistencies noted, the raw values for Mar. 10 – 24, 2021 were not used but were replaced by linearly interpolated values utilizing the values of Mar. 9 and 25, 2021 as the basis for the interpolation. **IV)** When taking the daily values and summing over all age groups, the resulting values did not exactly match the PCR values in the file “*Test_pos_over_time.csv*” from Ref. [13]. A close examination of Fig. 9, shows that the two values do not line up and that the summed values are about 2 days later. The Ref. [13] values refer to the PCR sampling date, while the summed values are test results “reported by 8 a.m.” and thus evidently don’t correspond to the sampling date. While the values when shifted by 2 days still do not exactly match (probably largely from results reported later than 2 days after sampling), Fig. 9 shows that a 2-day shift provides a good overall match. Thus, from this point forward all the PCR age fraction results will have the needed 2-day shift built in so that they can be used with the PCR sampling date values that are the time basis of most of calculations and results reported herein. **V)** There were no daily AGE-based values available for the first few weeks of the pandemic with only a single integral AGE-based set of values covering that entire time period existing. The daily values were estimated using that single set of AGE-based fractional values multiplied by the daily PCR values from Ref [13].

**Fig. 9:**
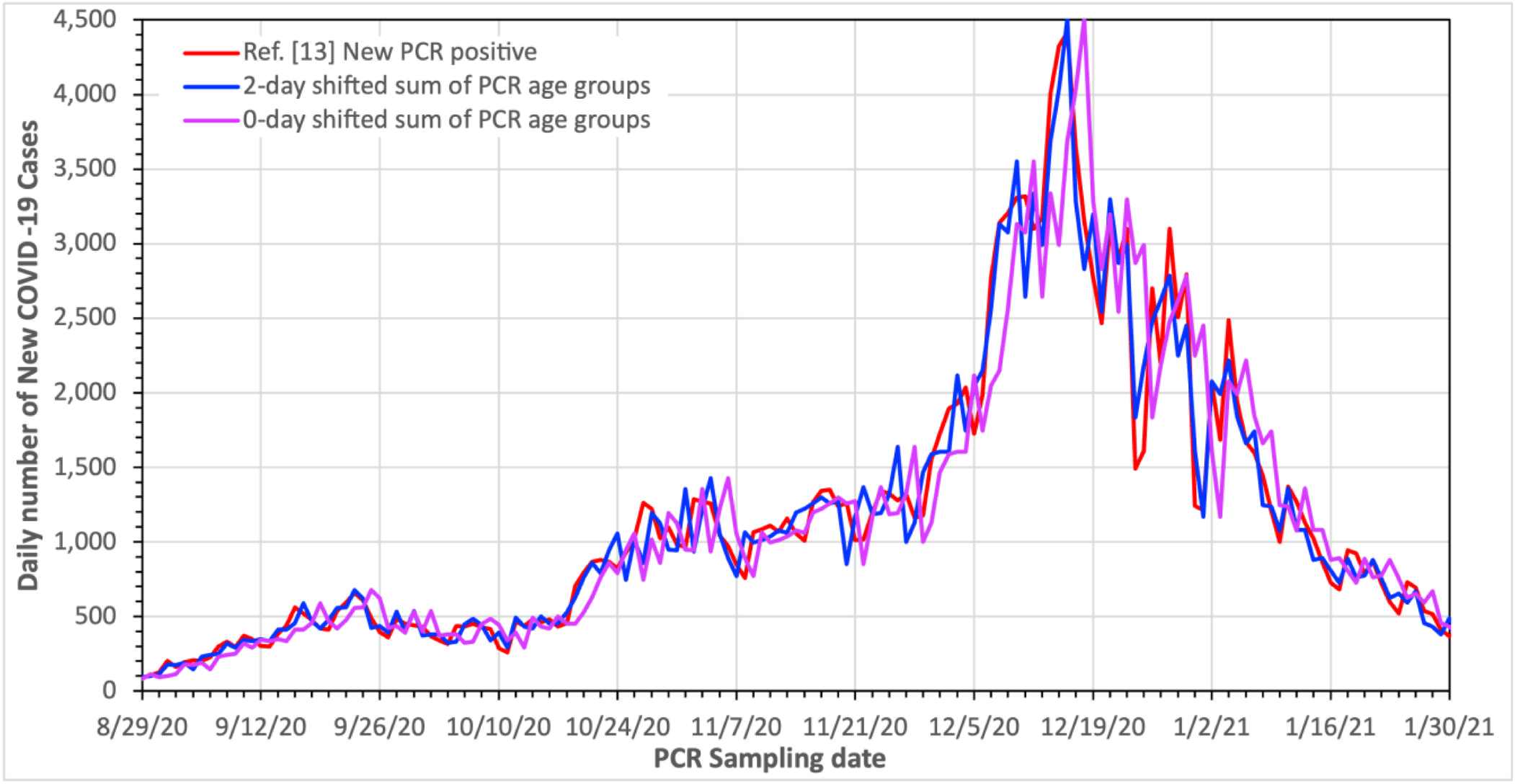
Daily values of the number of new COVID-19 cases in Denmark from PCR tests using Ref. [13] (red) and the sum over all age groups (magenta). Also shown are the sum over all age groups values but shifted 2 days earlier (blue), which illustrates the need for the shift. Horizontal axis tick marks are every 2 days. See point IV).

The Simple Method applied relies on the fact that over an “extended time period” from early Sept. into early Dec. 2020, the “Alpha variant” only adjusted *IDE* values were fairly constant, as were the fraction of COVID-19 PCR cases in the ≥ 60-year-old age group. The latter point can be seen in Fig. 10 where the fractions of daily PCR identified cases in each 10-year age group are shown as a function of time. Also shown is the fraction of cases in the ≥ 60-year-old age group. The rough constancy of the “Alpha variant” only adjusted *IDE* values in the “extended time period” can be seen in Fig. 7. In particular, the time period Sept. 9, 2020 through Dec. 8, 2020 inclusive (an integral number of whole weeks) was used as the “extended time period”. For this “extended time period” the average of the daily values of the ≥ 60-year-old fraction of new PCR cases was calculated. Likewise on a daily basis, the “7-day running average” of the daily value of the fraction of new PCR cases for those of age ≥ 60 years to the total number of new PCR cases was calculated. The “7-day running average” fraction for day τ actually used was in fact:

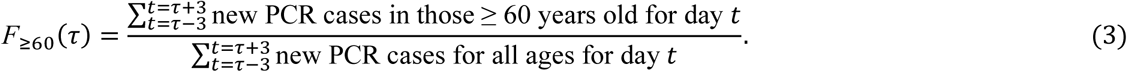

**Fig. 10:**
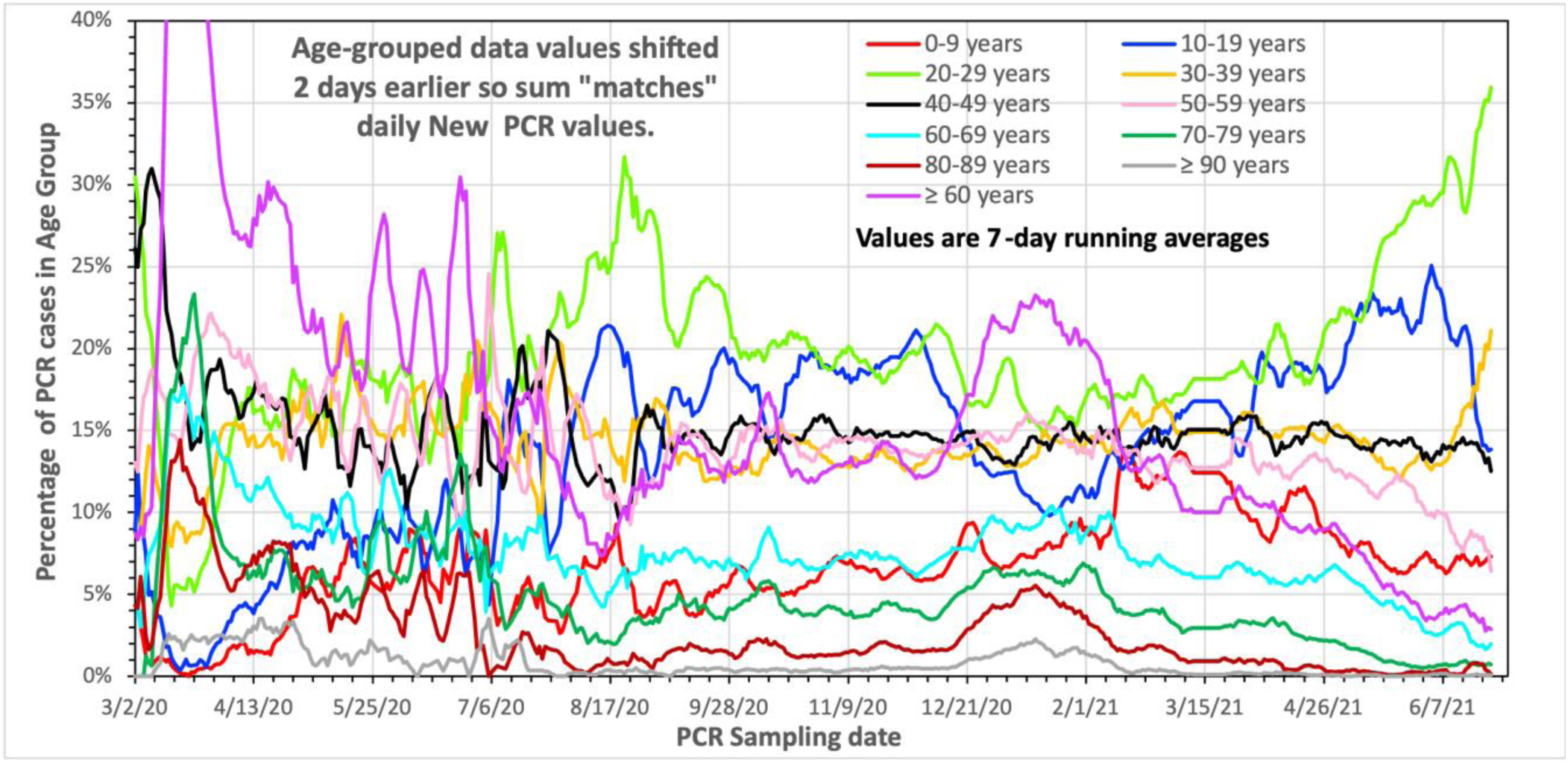
The daily 7-day “running average” values of the fraction of PCR confirmed COVID-19 cases in the different 10-year age groups and the ≥ 60-year-old age group. The 7-day “running average” values are actually the 7-day sum of the numerator values divided by the 7-day sum of the denominator values as in Eq. (3). Horizontal axis tick marks are at 1-week intervals. The values shown have issues **I)** – **V)** in the text addressed.

Likewise, the “average value” of the above for the “extended period” was defined and calculated as

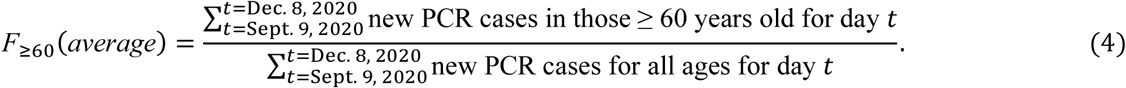

For the Simple Method, for any given date *τ*, the AGE and “Alpha variant” adjusted daily new hospital admission value is the daily new “Alpha variant” adjusted admission value for that day <u>multiplied by</u> the <u>ratio</u> of the “average value” of the daily new PCR fraction for those ≥ 60 years of age for the “extended period” to the daily “7-day running average” of the daily new PCR fraction for those ≥ 60 years of age for date *τ*, namely

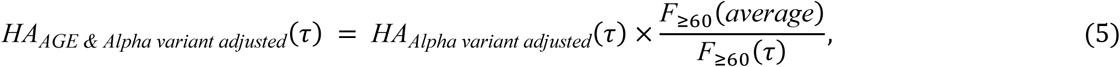

where, just as in Section 3.2, *HA*(*τ*) is the time shifted value of the hospital admissions on day *τ* with the subscript indicated adjustment.

It can be verified that the adjustment factor [*i.e.*, the ratio on the RHS of Eq. (5)] has the correct “polarity” as follows. When the fraction of COVID-19 cases in those aged 60 years or older is larger than normal or average, then the ratio in Eq. (5) will be < 1. This results in the AGE adjusted value of the new hospital admission being less than the unadjusted value, which is as it should be since otherwise the true number of COVID-19 cases derived from the hospital admission values would be too large. Likewise, when the fraction of COVID-19 cases in those aged 60 years or older is smaller than normal, then the ratio in Eq. (5) will be > 1. This results in the AGE adjusted value of the new hospital admissions being greater than the unadjusted value, which is also as it should be.

The AGE and “Alpha variant” adjusted daily new hospital admission values calculated via the above Simple Method must still be scaled in order to turn them into daily new actual COVID-19 case numbers. This is done as previously described by integrating the AGE and “Alpha variant” adjusted daily new hospital admission values from the start of the pandemic through the date corresponding to the Round 4 Prevalence derived number of cases. The scaling factor is the number by which the integral AGE and “Alpha variant” adjusted daily new hospital admission integral value must be multiplied to yield the Round 4 Prevalence derived number of cases.

As per Eq. (1), the ratio of the (daily values of the new PCR confirmed cases) to the (AGE and “Alpha variant” adjusted daily actual number of new COVID-19 cases given by the AGE and “Alpha variant” adjusted number of daily new hospital admission values multiplied by the above scaling factor) yields the AGE and “Alpha variant” adjusted daily *IDE* values. For comparison purposes, AGE and “Alpha variant” adjusted daily *IDE* values were also calculated as above, but using either ≥ 50-year_old PCR age fraction values or ≥ 70-year_old PCR age fraction values. The results are shown in Fig. 11 along with the “Alpha variant” only *IDE* results shown earlier in Fig. 7.

**Fig. 11:**
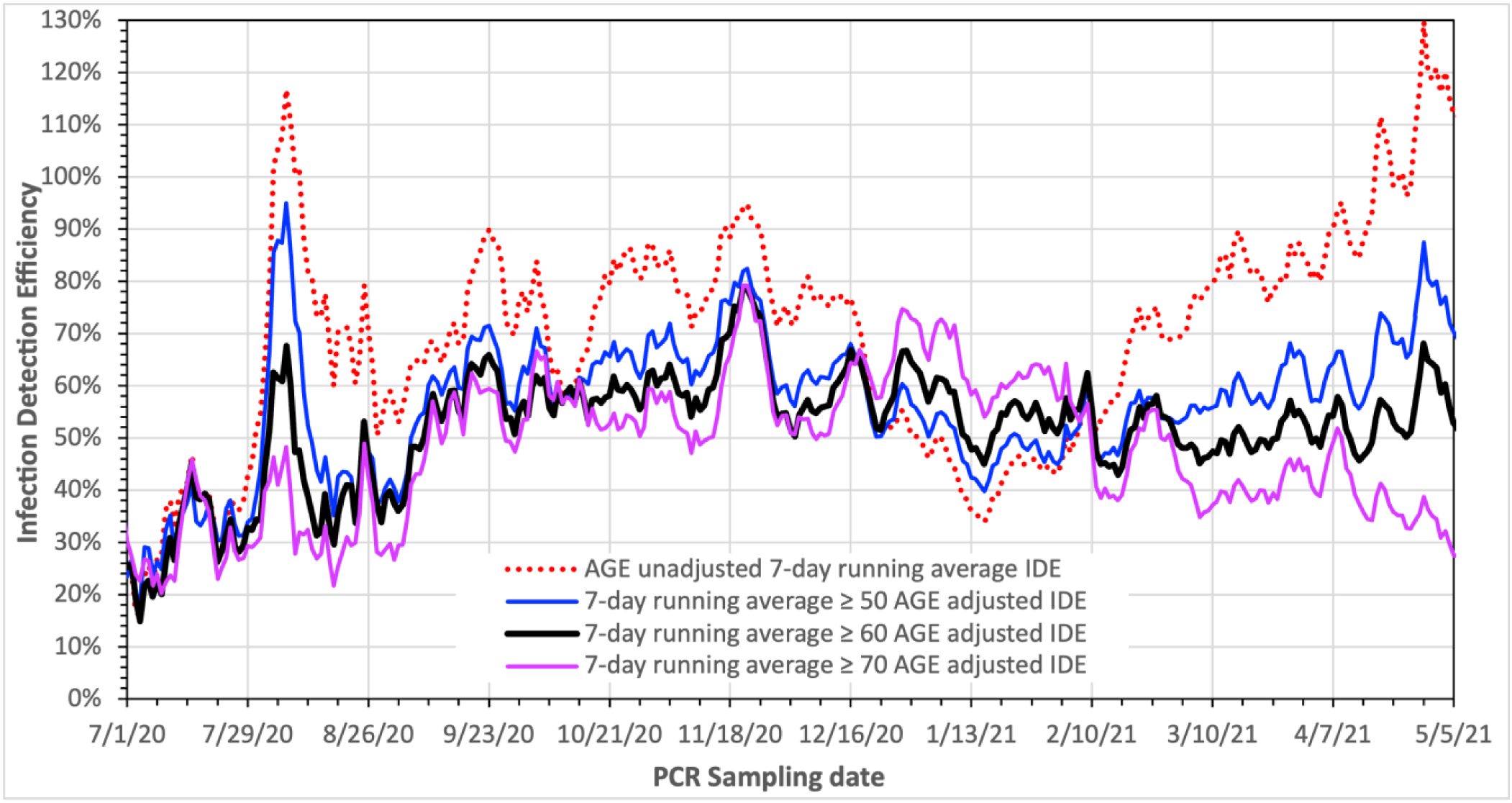
The daily 7-day “running average” of the AGE and “Alpha variant” Simple Method adjusted IDE values using 3 different age group bases. For comparison, also shown is the “Alpha Variant” only adjusted IDE result (the dashed red curve). Horizontal axis tick marks are at 1-week intervals. We believe that the ≥ 60 AGE adjustment (black curve) yields the best Simple AGE-Based Adjusted IDE results; see text for details.

The results which also include AGE adjustments show smaller variations in the *IDE* values than the “Alpha variant” only adjusted result. Overall, the ≥ 60-year-old based adjustment shows the steadiest results with an average value of ∼ 56% from mid-Sept. 2020 into early May of 2021, which using the 95% CI for the scaling factor range from Section 3.3 has a corresponding range of 53.0% to 59.4%. Also, including the AGE adjustments keeps the Aarhus outbreak values below 70%, almost fully removes the large dip from mid-Dec. 2020 to late-Feb. 2021, and largely overcomes the vaccination impacts. The ≥ 50-year-old based adjustment and ≥ 70-year-old based adjustment don’t do as well, seemingly overcorrecting in some locations and undercorrecting in other locations.

If one ignores the Aarhus related peak, during the time period from the start of Fig. 11 and extending into early Sept. 2020 it appears as though the integrated testing and contact tracing program was still ramping up as indicated by the predominantly increasing *IDE* values throughout that period. In addition to the highly focused contact tracing and intervention effort associated with the Aarhus outbreak peak, there is also now a clear peak in mid-Nov. 2020 where the AGE-based adjusted versions of the *IDE* reach about 80%.

### 3.8 Another Adjustment Approach

With the apparent improvement in the *IDE* values realized in the previous subsection, AGE-based adjustment methods were examined in greater detail. It was found that if the individual hospitalization fractions for the COVID cases in each age group are known, the AGE-based adjustments could be properly calculated to estimate the daily number of new cases based on the daily new COVID-19 “Alpha variant” only adjusted hospital admission values.

The proper approach requires the daily values of the fraction of new PCR verified COVID-19 cases sorted by age group. These fractional values are taken to vary day-by-day. In addition, for each age group, the fraction of COVID-19 cases that result in hospitalization is required. Unfortunately, these latter values do not seem to be available on a daily basis for the period covered in this paper. However, some integral values were found. In particular, we use integral values from two references. The first of these, [36], reports integral values for Denmark through May 19, 2020, while [40] reports values for Austria though Dec. 31, 2021. The raw values (columns ***B***, ***C*** and ***F***, ***G***, respectively) are given in Table 3 along with the values calculated from them (columns ***D***, ***E*** and ***H***, ***I***, respectively), which in turn are used to yield the ratio of the Danish to Austrian values (columns ***J*** and ***K***).

**Table 3.**
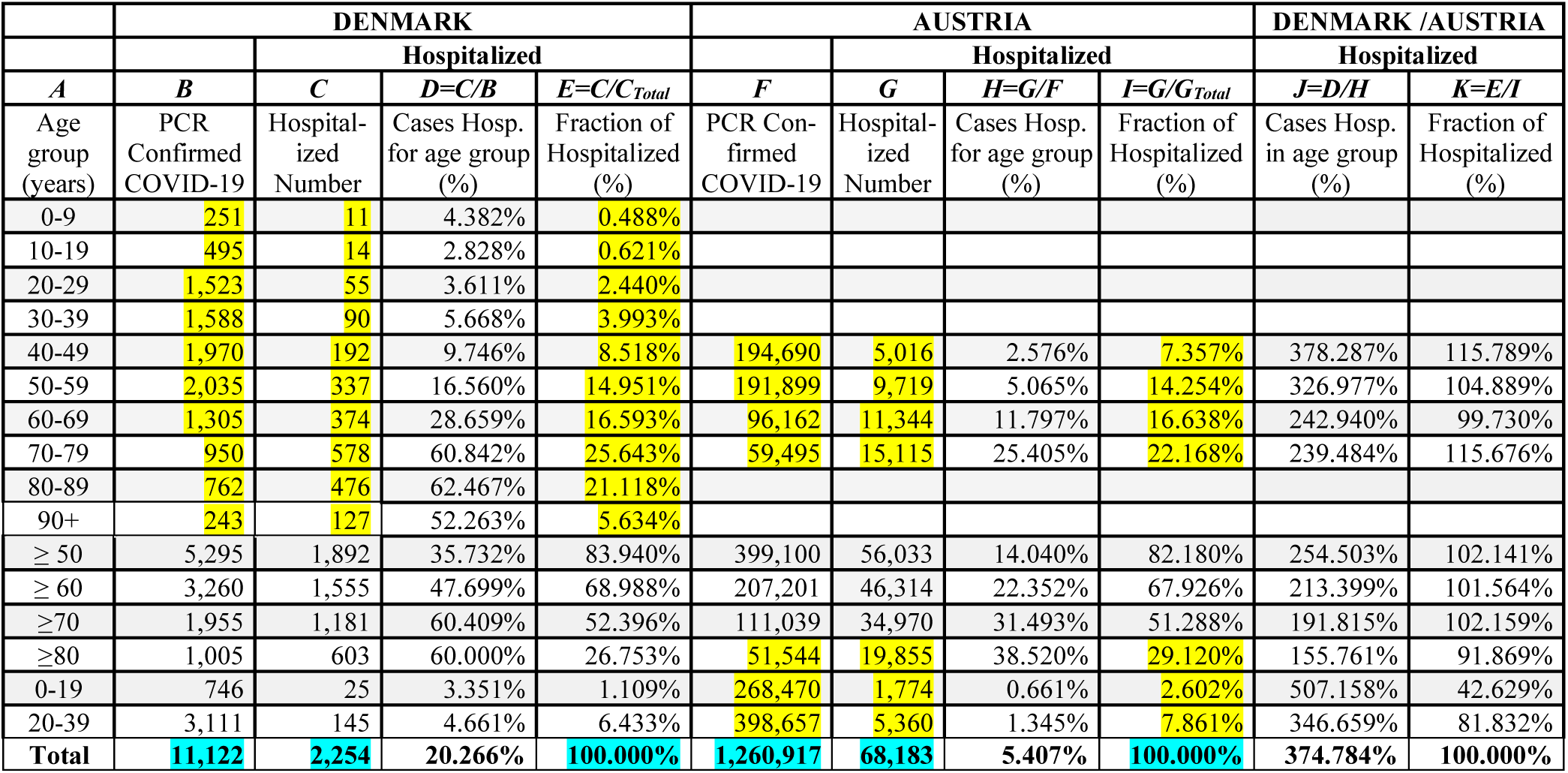
The PCR confirmed COVID-19 cases, resulting hospitalizations, and fractions by age for Denmark and Austria. The yellow highlighted values are from Refs. [36, 40], with the other values being calculated therefrom. The yellow highlighted entries are the ones summed to give the cyan highlighted “**Total”** values in the last row.

As daily values are not available, AGE-based hospitalization fractional values are assumed to be independent of time. Whether this is an appropriate assumption is of course an appropriate question. As no significant at home treatments for COVID-19 existed during the time period covered herein, to first order one would expect time independence. Also, hospitalization rate changes that could be attributed to COVID-19 variant effects are already adjusted for in the “Alpha variant” adjustments. However, the Danish and Austrian hospitalization rate values (col. **D** and **H**) are very different as seen in Table 3. Part of this difference can be attributed to the fact that the Danish values are from early in the pandemic when testing capabilities were insufficient to deal with all the cases, and milder illnesses were not tested. This meant that only the more serious cases were favored in testing which resulted in high hospitalization fractions. In contrast, the Austrian values sampled much later into the pandemic when there was sufficient testing capacity to deal with all the testing requests (including mild cases which were unlikely to require hospitalization). There is however a reservation with the Austrian values as they include later variants that were not properly adjusted for herein. Furthermore, the values also extend well into the vaccination time frame.

Despite the issues identified above, the situation is better than it seems. Although the AGE-based hospitalization fractions for Austria and Denmark typically differ by a factor of a few (see col. ***J***), it should be recalled that the fractional values are taken to be constant in time. Furthermore, after the AGE adjustments are applied to the COVID-19 “Alpha variant” adjusted hospital admission values, the integral of the resulting adjusted admission values is scaled to match the prevalence results. The different AGE adjusted hospital admission values using the Danish vs. Austrian hospitalization fractions will result in different scaling factors, which, when applied, will lead to similar *IDE* values provided that the “Fraction of Hospitalized” for Austria vs. Denmark are essentially the same. That they are is seen by comparing columns ***E*** and ***I*** in Table 3 or examining column ***K*** where their ratio values are essentially 100%, especially for the more elderly age groups, which dominate the hospital admissions.

The proper method of adjusting the hospital admission values for AGE-based effects follows. First, we define several variables that will be used. Let *PCR_j_* (*t*) be the number of new PCR confirmed cases for day *t* and age group *j*, there being *n* age groups in total. The fraction of new PCR confirmed cases for day *t* and age group *j* is defined by

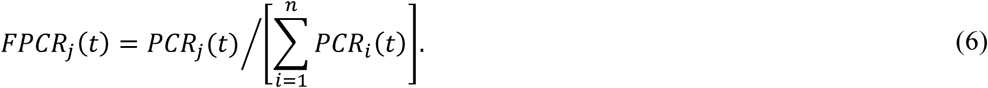

The fraction of COVID-19 cases for age group *j* that end up being hospitalized is defined by

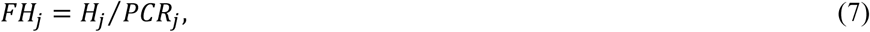

where *PCR_j_* is the integral number over time of PCR confirmed cases for age group *j* and *H_j_* is the integral number of those PCR confirmed cases for age group *j* that end up being hospitalized due to COVID-19, these values being given in Table 3 for Denmark and Austria respectively from Refs. [36,40]. Note again that these values are taken to remain constant over time.

Prior to the application of the prevalence scaling factor, the correction factor [*CF*(*t*)] to adjust for the age-effects associated with the COVID-19 new daily hospital admission values to turn them into actual COVID-19 case values is given by

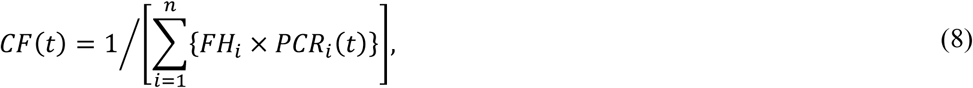

where the sum is over all age groups *n*. The AGE and “Alpha variant” adjusted hospital admission values are gotten by multiplying the daily new “Alpha variant” adjusted hospital admission values derived in Section 3.2 by the adjustment factor of the appropriate date given by Eq. (8). Next, the integral of these AGE and “Alpha variant” adjusted hospital admission values through the date corresponding to the Round 4 Prevalence case value is calculated. Then, using the same procedure as in Section 3.3 (but this time using the integral of the current AGE and “Alpha variant” adjusted hospital admission values), a new scaling factor to convert the AGE and “Alpha variant” adjusted hospital admission values into actual COVID-19 cases is calculated and then applied. With the resulting revised actual COVID-19 case numbers now in hand, revised *IDE* values can be calculated using Eq. (1). In performing the various calculations mentioned in this subsection it is important to note that time shifts are needed in order to get the proper results. These time shifts were mentioned in Sections 3.1 and 3.7. Also as mentioned earlier, where fractions are involved, 7-day running averages are calculated for the numerators and denominators prior to calculating the fraction values rather than using the 7-day running averages of the daily fractional values. The AGE and “Alpha variant” adjusted versions of the *IDE* values calculated using the Danish and the Austrian Age-based hospitalization fractions are plotted in Fig. 12 along with the earlier AGE unadjusted and the ≥ 60-year AGE adjusted values previously shown in Fig. 11 for comparison. A comparative evaluation of the performance of the different approaches will be provided later in Section 4.

**Fig. 12:**
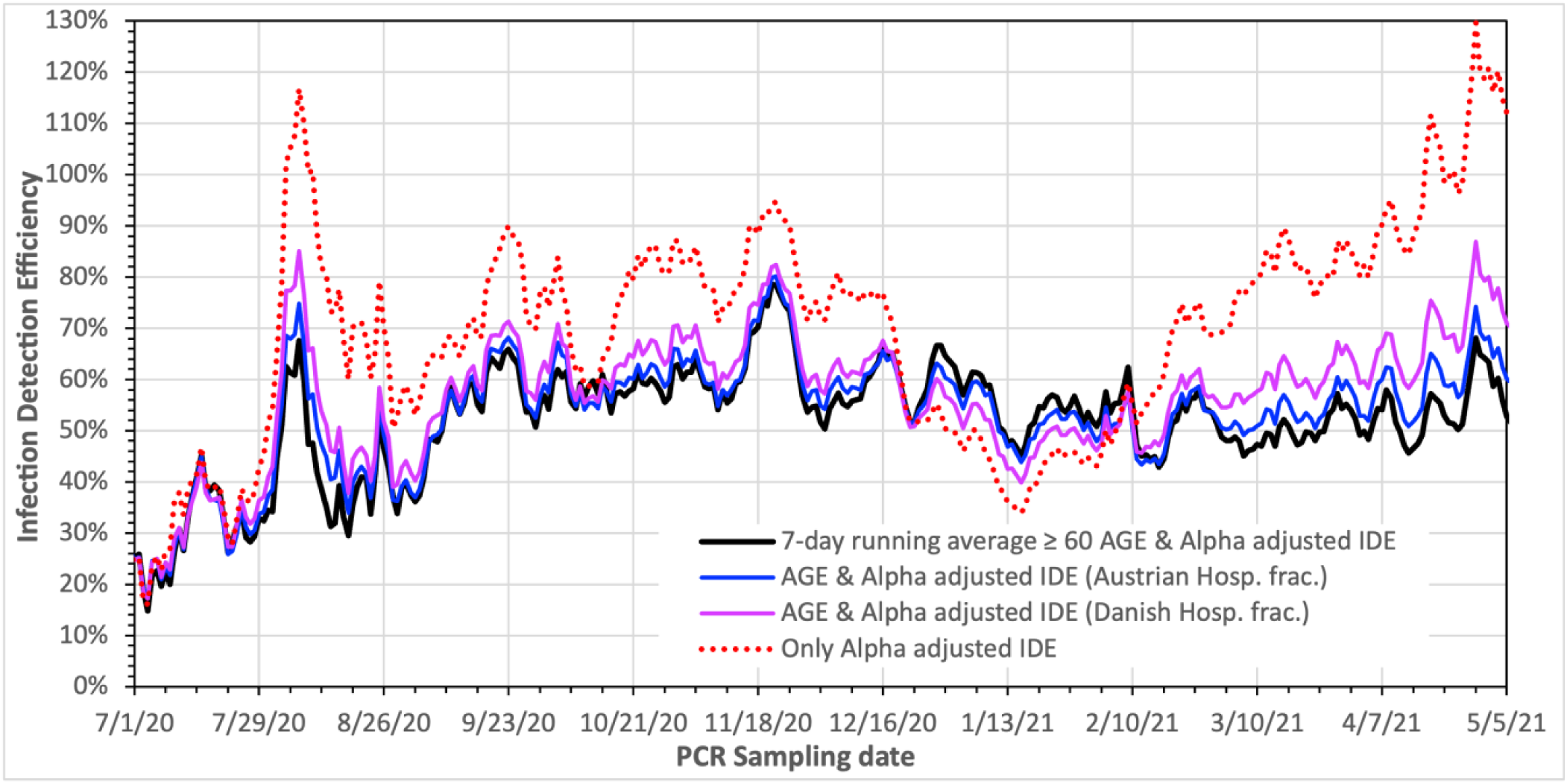
The daily 7-day “running average” of the AGE and “Alpha variant” adjusted IDE values using the Danish and Austria AGE-grouped hospitalization fraction values. For comparison, both the AGE and “Alpha variant” adjusted IDE values using the ≥ 60-year-old age based Simple Method (heavy black curve) and the “Alpha Variant” only (dashed red curve) adjusted IDE results are also shown. Horizontal axis tick marks are at 1-week intervals.

### 3.9 Exploration of Other Hospitalization Delays

As noted in Section 3.1, the time that serious COVID-19 cases end up being hospitalized is a bit uncertain. For the results presented up to this point in the paper, hospitalization was assumed to take place 5 days after COVID-19 symptoms appeared in those that would require hospitalization. This was the “best” value based on the references given in Section 3.1. Several other times of hospitalization were explored to verify this. To be more specific, the timing values previously adopted were: exposure to COVID-19 virus on day 0; symptoms (if they occur) on day 6, hospitalization (if it occurs) on day 11; and antibodies first detectable in prevalence measurements on day 14. The other hospitalization timing cases considered all also have the same values for exposure, symptoms, and antibody detection, but have hospitalization occurring on day 6 (0-day shift), day 8 (2-day shift), day 10 (4-day shift), and day 13 (7-day shift) relative to the symptom & PCR times. For these cases, revised *IDE* values using the Simple Method are calculated and displayed in Fig. 13, along with the “standard” value which uses day 11 (5-day shift). It should be noted the changing the hospitalization delay impacts the “Alpha variant” adjustment for hospitalization values, the scaling factor for converting hospitalization values into COVID case values, and the AGE adjustment applied to the *IDE* values, although the impacts are not that large. In addition to changes in the “standard” *IDE* values, feature locations are slightly moved in time.

**Fig. 13:**
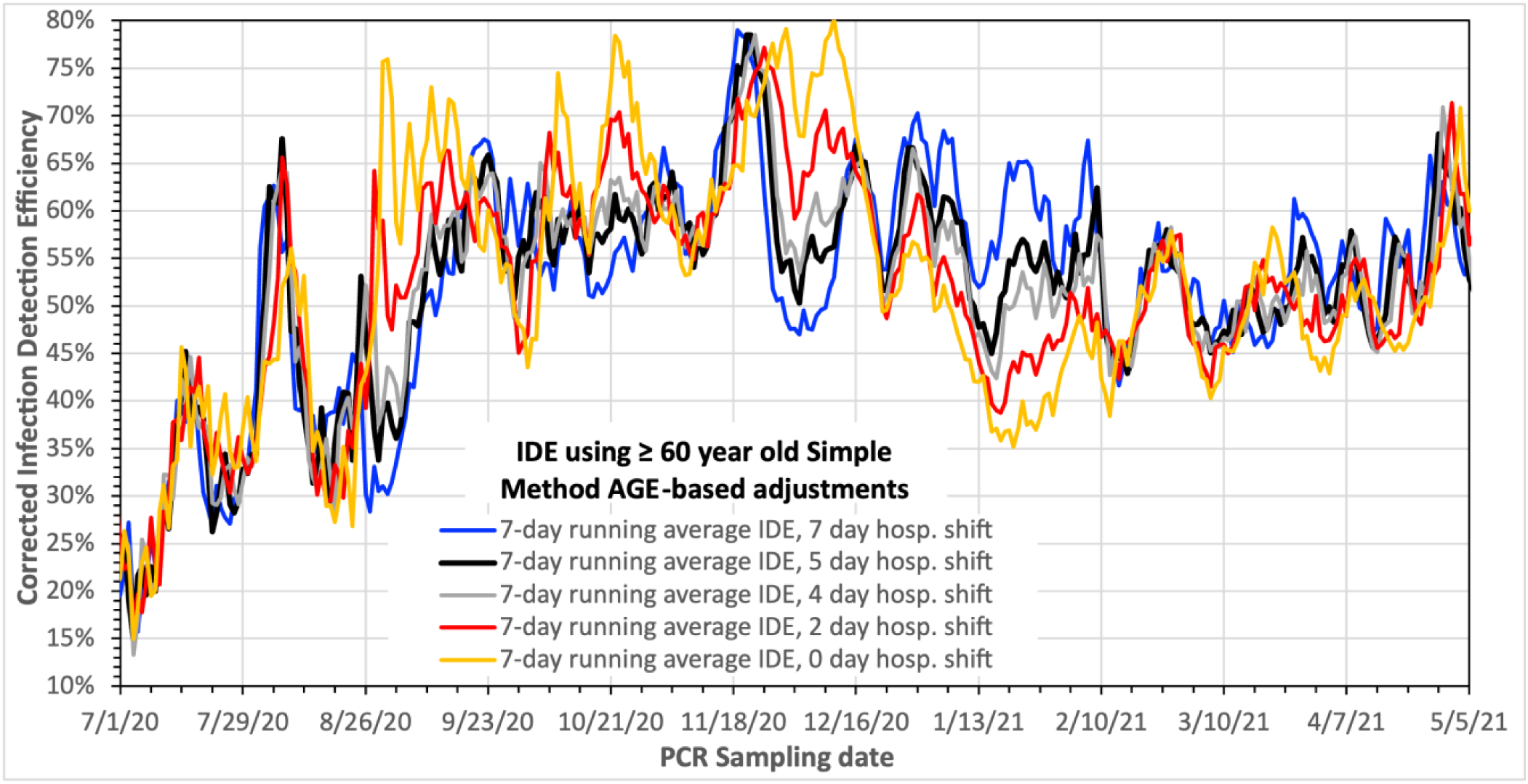
The “Alpha variant” and AGE adjusted 7-day “running average” of the daily values of the IDE for various different delays between when symptoms typically appear (day 6) and when hospitalization would occur if it is needed. The “standard” delay used in this paper is 5 days (**black curve**) (hospitalization on day 11; exposure on day 0). Based on the minimal variation between dip and peak values the “best” delay is either a 4 – or 5 – day delay reaffirming the originally chosen delay. Horizontal axis tick marks are at 1-week intervals.

Inspection of Fig. 13 confirms our original choice of the shift between when hospitalization occurs relative to when symptoms would first appear (same as PCR sampling date) is either 4 or 5 days. This is based on which curve(s) show the minimal variation between the dips and bumps in *IDE* values throughout the time period plotted. The 4 and 5-day shift curves perform about equally well.

## 4. Insights, policy implications, and questions

Based on the *IDE* values derived in Section 3, here we present insights gained by comparing these values with other measures of the pandemic. This provides a means of evaluating the effectiveness of the contact tracing and the testing strategies employed. We also examine the costs associated with policies Denmark adopted to deal with the pandemic and the performance of Denmark’s efforts relative to those of other countries. From these studies we delve into questions related to the implications for dealing with future pandemics.

### 4.1 *IDE* expectations

Before attempting to determine which of the combined AGE and “Alpha variant” adjusted *IDE* value sets developed in Sections 3.7 and 3.8 is “best”, the expected behavior of the *IDE* values is reviewed. COVID-19 cases are either symptomatic or asymptomatic. These two categories will have different impacts on the *IDE* value. We implicitly assume in the following analysis that the fraction of COVID-19 cases that are symptomatic vs. asymptomatic does not change over the course of the pandemic covered here.

Symptomatic cases as their name suggests are all assumed to yield noticeable symptoms typical of a COVID-19 infection. The symptoms will let those individuals know that they are ill and as a result these individuals are all assumed to “naturally” (i.e. without government intervention) get PCR tested to confirm that they either have COVID or that they are COVID free and instead have some other (non-COVID) infection. As a result of this “natural” behavior, all symptomatic COVID-19 infections will be caught. This will be the case regardless of governmental interventions such as forced lockdowns or reopenings of society or government required testing. Also, required testing before being allowed to travel, to return to work, to return to school, to attend large events, etc. will not change the presumption that all symptomatic cases will be caught. Based on the above and the definition of the *IDE* given by eq. (1) as being the daily ratio of the number of New PCR confirmed COVID-19 cases to the actual number of New COVID-19 cases results in the *IDE* value due to symptomatic cases being constant, at least once there is sufficient PCR testing capacity to meet demand.

The symptomatic COVID-19 individuals are assumed to reach out to their acquaintances to let them know that they were likely exposed and thus should get PCR tested. (We assume that such outreach does not change with time.) The acquaintances who test positive will also fall into either the symptomatic category or the asymptomatic category. The symptomatic acquaintances would have been caught regardless of the outreach and thus have won’t change *IDE* values. However, the asymptomatic acquaintances would not have been caught without the outreach. Thus, the outreach results in an increase in the *IDE* value. However, given that such outreach is assumed not to change and is directly tied to the number of symptomatic cases that resulted in the outreach, the size of the increase will not change as a result of any governmental interventions.

In addition to the outreach to acquaintances that happens spontaneously, the STPS will identify close contacts of the symptomatic COVID-19 cases and let them know that they need to get PCR tested. As a result of the follow-on testing of the close contacts, the positive PCR identified individuals found will be subdivided into the symptomatic or asymptomatic categories. As always, the PCR positive symptomatic close contacts will be caught regardless and thus won’t change the constant *IDE* value. However, the asymptomatic close contacts wouldn’t have been caught without the STPS contact tracing. Thus, these now caught asymptomatic close contact cases will also result in an increase in the *IDE* value. However, just as for the outreach cases in the previous paragraph, the close contact cases are all directly related to the initial symptomatic cases. Thus again, while the overall *IDE* value will be higher as a result, the size of the increase will remain constant in spite of interventions. The exception to this will be if the contact tracing program itself changes which will result in fractionally more or fewer asymptomatic cases being caught which would in turn change the *IDE* value in time.

One final point regarding the above discussed second generation symptomatic cases resulting from outreach to acquaintances or the contact tracing effort is that these cases might be caught sooner than if the outreach or contact tracing had not occurred. This is because they might now be tested during the couple of days when the cases are PCR detectable, but don’t yet display symptoms. The impact of this point (if any) on the *IDE* values is expected to be quite small. First of all, the number of cases involved is expected to itself to be quite small. Furthermore, New PCR cases can only be caught once; namely after initially being caught, they will now longer be New if caught again. However, on the date that they would have been caught without outreach or close contact tracing the cases will likely be filled in by other cases caught early as a result of outreach or close contact tracing. Thus, the net impact on the *IDE* would be zero and therefore we simply ignore this final point in terms of *IDE* values.

To summarize all of the above, it is expected that once there is sufficient testing to meet demand, the symptomatic COVID-19 cases will yield an unchanging contribution to the daily *IDE* values regardless of interventions by the government or other entities. The only exception to this would be due to changes in the effectiveness of the STPS close contact tracing effort and would only apply to asymptomatic cases flagged by that effort on the previously identified symptomatic population.

This then only leaves asymptomatic cases (other than those above that were directly linkable to symptomatic cases as mentioned above) available to cause changes to the daily *IDE* values over time. It is assumed that these infected asymptomatic individuals would be essentially randomly distributed amongst the population and being asymptomatic would have no reason to come in to be tested unless “forced”. Such “forcing” could be very mild such as government implied patriotic duty to be tested or more vigorous “forcing” such as testing being required prior to travel, etc., as mentioned previously, or severe “forcing” such as being quarantined or fined until tested.

With the asymptomatic cases mentioned in the preceding paragraph being effectively randomly spread throughout the Danish population, without well targeted testing, only large-scale testing of the Danish population would be capable of finding them in significant numbers. As of mid-Sept. 2020, about 0.8% of the Danish population was being tested per day (see upper plot of Fig. 14) and by the end of our period of study (early May 2021) ∼3.0% of the Danish population was PCR tested per day. In between these 2 times, there is peak value of ∼2.3% in the tests/person or equivalently the percentage of the Danish population tested per day on Dec. 19, 2020. This peak is associated with the peak in the large number of COVID cases, at that time as well as required testing in order to travel for the Christmas holidays. One might expect that the daily fraction of the asymptomatic cases caught to be equal to the fraction of the Danish population tested on a daily basis, however it is likely to be several times larger. The reason for this is because infected individuals will yield positive PCR tests for a number of days. This period will be approximately the same as the period of time during which people are contagious, which for the purposes of the following estimates we take to nominally be 10 days. As such, if the testing is random in nature, then to first order, testing *N* % of the Danish population in a single day would catch 10*N* % of the asymptomatic cases in that day of testing. Thus, if one takes the above value of 0.8% of the population being tested per day after the 1^st^ week of Dec. 2020, the one would catch 8% of the asymptomatic cases, while the testing 2.3% of the population per day with the Dec. 2020 peak would catch 23% of the asymptomatic cases, and testing 3.0% of the population per day in early May 2021, would catch 30% of the asymptomatic cases. However, if this testing is caried on for multiple days, the actual integral value will be smaller due to second order effects such as randomly testing some people multiple times within the interval, coupled with the fact that positively detected people can only be New cases once. This would reduce the 8% value to ∼7.7%, the 23% value to 20.8%, and the 30% value to ∼26.3%.

**Fig. 14.**
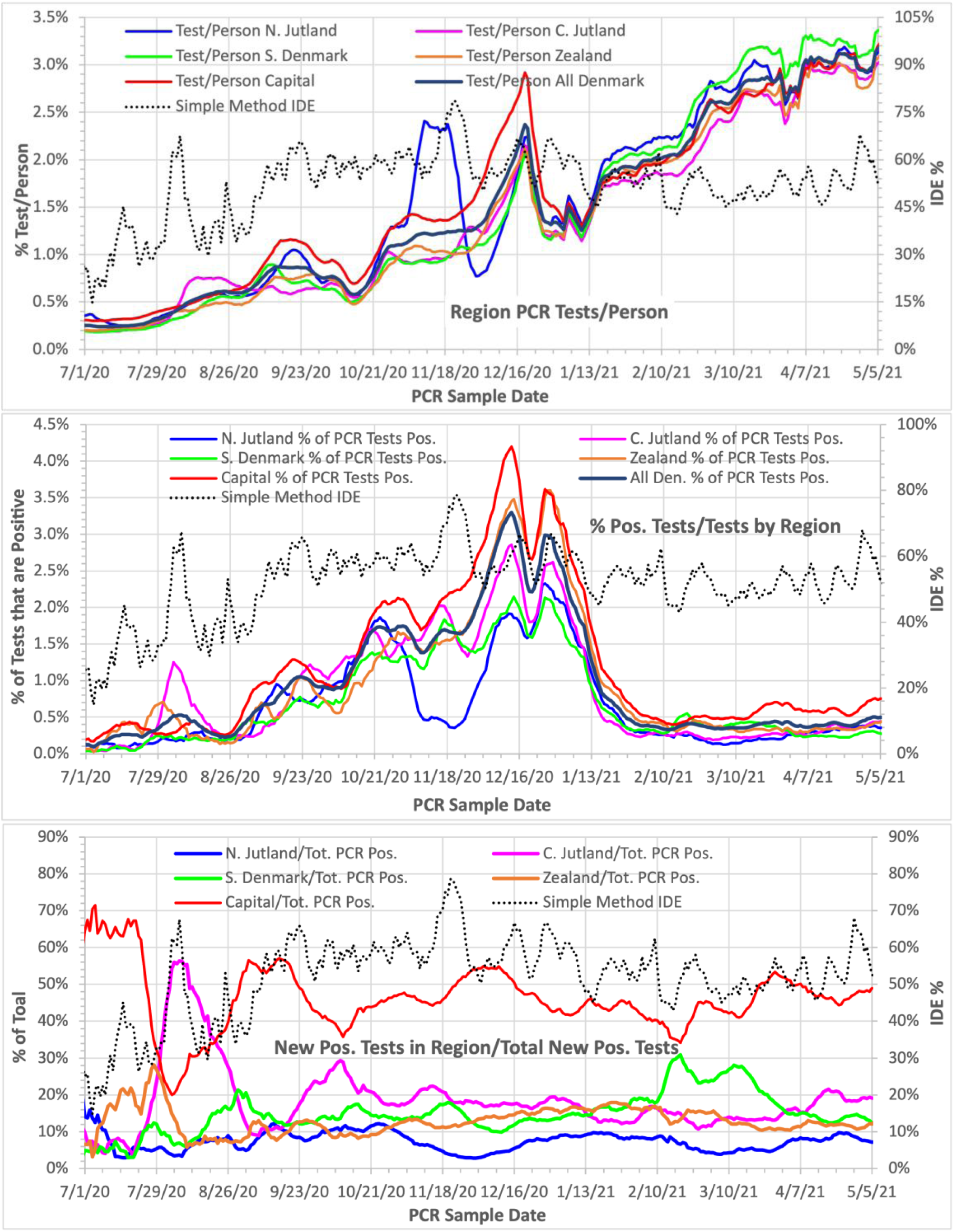
Upper plot: The fraction of Denmark regions’ population PCR tested on a daily basis (i.e. daily PCR tests/person). Middle plot: The ratio of the number of New positive PCR tests to the number PCR tests given on a daily basis for each region and all of Denmark. Lower plot: The ratio of New positive PCR tests in each region to the same for all of Denmark on a daily basis. For comparison, the dashed curves (right axes) give the IDE values. Horizontal axis tick marks are at 1-week intervals. “7-day running averages” as defined earlier are shown.

There are however a significant number of other complicating factors that make it very difficult if not impossible to determine reliable values for what fraction of the asymptomatic cases are realistically expected to be caught. Examples of the complicating factors are how random the testing actually is, what fraction of the testing is devoted to testing the same people repeatedly, the PCR test sensitivity decreasing with time into the infection, etc. These examples would further decrease the fraction of asymptomatic caught by large-scale testing.

Some other factors in contrast could increase the fraction of asymptomatic cases caught. Two specific examples are the very effective targeting of testing and the use of RA tests for COVID detection in addition to the PCR testing. While the number of RA tests/day carried out was completely negligible all the way into early 2021 (less than 10% of the number of PCR tests/day as of mid-Feb. 2021), thereafter their use rapidly expanded reaching values comparable to PCR test numbers by early April 2021, and were roughly double that by early May 2021. The RA tests are however not that effective as PCR tests since they have considerably lower sensitivity than the PCR tests. According to Ref. [41], broadly speaking and relative to PCR tests, RA tests have sensitivities of ∼75-85% for people with symptoms, but only ∼45–55% for the people without symptoms that are currently being considered here. (It is again noted that any positive RA tests results had to be confirmed by PCR tests in order to be counted as a case.) In spite of the issues concerning the sensitivity of RA tests for asymptomatic cases, whether the testing is randomly distributed amongst the population, etc., the significant increase in testing with time and its scale, especially in the April - May 2021 time frame, should lead to a readily evident increase in the capture of asymptomatic COVID-19 cases and hence a simultaneous overall increase in time of the *IDE* value. In addition, effective targeting of testing can also result in the capture of asymptomatic cases during the time period when it occurs leading to a coincident peak in the *IDE* value. The effective targeting also implies that the contact tracing is more effective, but not necessarily whether this is due to the tracking of individual cases vs. identifying specific groups that need to be heavily tested.

To summarize this subsection, to first order, (once there is sufficient testing capacity to meet demand) it is expected that the *IDE* value due to the symptomatic COVID-19 cases will remain constant, while any changes observed in *IDE* values will be due to changes in the ability to catch asymptomatic cases as a result of enhanced targeted testing or overall increasing testing levels once they have become quite large.

### 4.2 *IDE* and contact tracing/testing results and what they say

We now compare the expectations of the behavior of the *IDE* values discussed in the previous subsection with what was actually observed in Fig. 12 for the 3 different AGE and “Alpha variant” adjusted *IDE* variants (solid line curves) derived in Section 3.

As seen in Fig. 12 (with the exception of 2 significant peaks and ignoring the initial ramp up), relatively stable *IDE* values were found after both AGE and “Alpha variant” adjustments were applied. The initial ramp up was taken to be complete by Sept. 15, 2020. During the ramp up, a large *IDE* peak occurs in early Aug. 2020 which is associated with the early Aug. 2020 Aarhus COVID outbreak. After the ramp up was complete, apart from a second peak in the mid-Nov. 2020 time frame and relatively minor fluctuations, the *IDE* values remain relatively constant through the end of 2020. Also, none of the minor fluctuations could be definitively associated with distinct changes in policy or testing/contact tracing changes. During the first two months of 2021 the *IDE* values continue to also remain relatively constant although their overall value was slightly lower than the “*steady*” period of 2020. The 2^nd^ major shutdown was also gradually lifted starting Mar. 1, 2021 and was accompanied by a renewed emphasis on large scale testing. Starting towards the end of Mar. 2021, a continuing increase in the *IDE* values is seen for at least some of the AGE and “Alpha variant” adjusted *IDE* values. It is most evident for the “Danish Hosp. frac.” curve and least evident for the ≥ 60-year-old based Simple Method. (The “Only Alpha variant” curve is ignored as this adjustment alone was known to be insufficient to yield reliable COVID case numbers.)

Examining the *IDE* values during their “*steady*” period of 2020 [Sept. 15 through Dec. 31, 2020, but excluding the mid-Nov. 2020 peak period window (Nov. 11 – Nov. 29, 2020)], the Mean (*M*) and Standard Deviation (*SD*) values for each of the 3 solid curves in Fig. 12 were calculated. The corresponding % values for the AGE and “Alpha variant” adjusted versions are: ≥ 60 Simple Method, *M* = 58.72, *SD* = 3.98; Austrian, *M* = 59.84, *SD* = 3.96; and Danish, *M* = 62.27, *SD* = 4.95. As this “*steady*” period excludes both significant *IDE* peaks and any associated enhanced testing/contact tracing efforts, the “steady” period *IDE* values are considered to be those of “*normal*” testing and contact tracing efforts.

As noted earlier, following this “*steady*” period, the *IDE* values were slightly smaller during the first 2 months of 2021. The *IDE* % values drop to *M* = 53.42, *SD* = 4.95 for the ≥ 60 Simple Method; *M* = 51.80, *SD* = 4.60 for the Austrian hospitalization fraction adjustments; and *M* = 50.62, *SD* = 5.21 for the Danish hospitalization fraction adjustments. The root cause of this lowering of *IDE* values is unknown although it roughly coincides with the duration and time of the second lockdown starting Christmas day 2020 and ending Mar. 1, 2021. Finally, starting towards the end of Mar. 2021, the 3 AGE and “Alpha variant” adjusted versions start to diverge in behavior. Aside from the ongoing fluctuations, the 60-year-old based Simple Method on average has more or less constant average *IDE* values throughout the period while both Austrian and Danish hospitalization fraction-based adjustments yield increasing *IDE* values with time; the Danish hospitalization fraction version yielding the larger increase.

Concentrating on the above “*steady*” period ∼60% *IDE* value for now, we compare it to other information about the pandemic to gain insights about Denmark’s “*normal*” testing and contact tracing effort. In particular, we focus on the fraction of COVID-19 cases that are asymptomatic. The reported values for that fraction in Denmark has considerable variability. For instance:

A. The fraction of COVID-19 antibody prevalence positive subjects reporting no symptoms for Round 2 [sampling from weeks 34-36 (Aug. 17 – Sept. 6) of 2020] of prevalence testing in Denmark was 45% [23], while for Round 3 [sampling from weeks 38-51 (Sept. 14 – Dec. 20) of 2020] that fraction had decreased to only 27% [22]. The low values of “no symptom cases” for Round 3 is likely due to several factors. First, prevalence tests themselves provide no information as to when positive testing individuals actually had COVID nor whether their cases were asymptomatic or symptomatic. Instead, the information about symptoms was the result of questionnaires that covered the period from the start of the pandemic to the time of the prevalence test. As such, it is quite likely that a number of what were actually asymptomatic cases were falsely implied to be symptomatic. This is because at other times these people also had non-COVID respiratory infections that nonetheless caused COVID-like symptoms. This is also hinted at by the low fraction of PCR tests that are positive even relatively early on in the summer of 2020 when testing rates were relatively low as seen respectively in the middle and upper plots of Fig. 14. The probability of having such other infections increases with the duration of the time period from the start of the pandemic covered by the prevalence measurement. In addition, it further increases when it includes times with higher respiratory infection rates such as in the latter part of the calendar year into which Round 3 extends. Thus, one would expect that the higher “asymptomatic case” fraction seen for Round 2 (45%) to be closer to the truth. In fact, its value is close to the 41% asymptomatic fractional value from Ref. [2] reported in the Introduction.
B. An early review article [42] covering the period through late May 2020 reported that “Asymptomatic persons seem to account for approximately 40% to 45% of SARS-CoV-2 infections”. Another later review article [43] covering the period through very early Apr. 2021 which looked at over 350 studies reported that “the percentage of cases that were truly asymptomatic among laboratory-confirmed cases was 35.1% (95% CI: 30.7% to 39.9%”).

The asymptomatic case values reported in A) and B) are very close to the (100% – “*steady*” period *IDE* average values from Fig. 12); i.e. ∼ 40%. This implies that the testing/contact tracing program as “*normally*” implemented caught very few of the asymptomatic cases. If it had caught such cases, the *IDE* values would be substantially larger and hence the 100% – *IDE* average values would have been substantially smaller than the fraction of asymptomatic cases.

Thus, as typically implemented, the inability of the testing and contact tracing program to find a notable number of asymptomatic cases indicates that it was insufficient. As noted in Section 1.2, the number of contacts traced averaged between 2 and 3 per confirmed COVID-19 case. This number was on average either too small or done too late after infections began to prevent a further spread of cases (or perhaps both). While the effort initially appeared to maintain a relatively low number of COVID cases during the summer and early fall months of 2020, this was more likely due to higher air temperature and humidity as well as fewer contacts occurring in confined spaces due to longer daylight hours, more outdoor activity, and summer holidays away from schools [e.g. 44-47]. Later in the year as one moves towards winter, far more contacts of longer duration occurred within confined close quarters, while air temperature and humidity both dropped. In light of this, it is clear that the “*normal*” contact tracing program had virtually no ability to contain the then rapidly growing number of cases resulting in the need to implement increasingly restrictive policies and then another lockdown of Danish society at Christmas 2020.

This is not to say that aggressive and well-targeted contact tracing/testing efforts could not be effective in controlling the pandemic. In fact, highly focused contact tracing/testing efforts appeared to have been successful in at least one instance in Denmark, namely the effort associated with the Aug. 2020 Aarhus COVID outbreak. The upper plot in Fig. 14 shows a significant increase in number of tests per person in the Aarhus municipality during the time of the Aug. 2020 *IDE* peak. In addition, the middle plot of Fig 14. shows that the testing was well targeted as the fraction of tests done that came back positive show a very large increase (by a factor of 3 to 4) at the time of the *IDE* peak. One also notes that during this outbreak the local *IDE* value for the Aarhus municipality must have been significantly above the Fig. 12 black curve value of ∼ 60 to 65% for the country as a whole at that time. This is because the Aarhus outbreak only contributed up to 45% of the total COVID-19 cases in Denmark in that period as seen in Fig. 6. When this fact is coupled with the observation that the *IDE* for Denmark as a whole was still ramping up, the contact tracing/testing campaign effort in Aarhus must have resulted in an *IDE* value approaching 100%. An estimate of the actual Aarhus municipality value can be obtained as follows:

- If during the outbreak one takes the above value of 45% of Denmark’s cases being in the Aarhus municipality, this means that 55% of the cases were in the remainder of the country.
- From the Fig. 12 black curve immediately before and after the Aarhus peak, we see that average value of the *IDE* would have been ∼35% during the peak period had enhanced targeted localized contact tracing/testing effort in Aarhus not been implemented.
- The actual value of the *IDE* in the peak region was ∼ 63% as seen from the Fig. 12 black curve.
- Putting all this together yields 63% = 35% × 55% + *Actual IDE in the Aarhus municipality* × 45%
- Solving this yields *Actual IDE in the Aarhus municipality* = [63% – (35% × 55%)]/45% = 97.2%

This high local value for the *IDE* resulting from the enhanced targeted and localized contact tracing/testing effort when combined with local contact restrictions was enough to bring the Aarhus outbreak under control as seen in Fig. 6. However, with the local value being nearly 100% [meaning that essentially all COVID cases (including the asymptomatic ones) were caught], the enhanced targeted and localized contact tracing/testing effort by itself (i.e., without local contact restrictions) should have been sufficient to bring the outbreak under control.

The other peak in the *IDE* in Fig. 12 which is seen in the mid-to-late Nov. 2020 time frame has the *IDE* rise from its typical value of ∼60% to just under 80%. This is also the time frame when an increased number of efforts were underway to deal with the mink COVID-19 outbreak in the North Jutland region of Denmark. The efforts included a lockdown and a large increase of testing which is seen in the upper plot of Fig. 14 for the North Jutland region. The testing peak however starts before the *IDE* peak begins and the testing starts to decline just as the *IDE* values actually peak. Also, the testing seems to be very poorly targeted as the middle plot of Fig.14 shows a dip at the time of the *IDE* peak. In fact, the tests per person values and *IDE* values seem to be completely out of phase. Another action taken during the Nov. time frame was the culling of the entire mink population of Denmark. Given all of these efforts, there is a natural tendency to attribute this mid-to-late Nov. 2020 *IDE* peak to the enhanced efforts in North Jutland. However, if one examines the lower plot of Fig. 14 one sees that in the associated time frame of the Nov. 2020 *IDE* peak, the efforts in North Jutland cannot possibly explain the increase in the *IDE* values for the whole country. With only 5% or less of the total cases in Denmark being in North Jutland at that time (and decreasing), there are simply nowhere close to enough COVID-19 cases in North Jutland to cause an increase of the *IDE* for the whole country from ∼60% to ∼80%.

The only other changes seen in the mid-to-late Nov. time frame in Fig. 14 that could possibly be associated with the peak in *IDE* values are small peaks in the middle plot for Central Jutland and Southern Denmark and perhaps coupled with the rise in the fraction of tests that were positive in the Capital region. It is also noted that there were increasing levels of mink related COVID-19 cases in the Central Jutland and Southern Denmark regions at that time. Whether or not these small peaks and/or the Capital region rise are associated with the *IDE* peak is however uncertain. We are also unaware of enhanced testing/contact tracing efforts in those regions at that time. Thus, unlike the case of the Aarhus peak we are unable to make a definitive association of the mid-Nov. 2020 *IDE* peak with an enhanced and targeted testing and contact tracing effort.

Looking more closely at the impacts of increased levels of testing on the *IDE* values, the predictions made in section 4.1 either don’t show up or are smaller than predicted for the daily testing values found. In the upper plot of Fig. 14, one sees that the black curve which is the percentage of the Danish population tested on a daily basis rises from about 0.8% at the start of the “*steady*” period to about 2.3% just before Christmas 2020 and shortly before the end of the “*steady*” period. Going back to the predictions of section 4.1, with a 10-day detectability window and a 40% value for the fraction of asymptomatic cases, the fraction of asymptomatic cases caught should rise from ∼7.7% at the start of the “*steady*” period to ∼20.8% at the pre-Christmas 2020 testing peak. Subtracting these two values one is left with a predicted increase of 13.1% in the fraction of asymptomatic cases caught by the increase in testing, assuming it was random. Taking into account that ∼40% of the cases are assumed to be asymptomatic, in terms of actual cases, the *IDE* values should increase from 63.1% (i.e., 60% + 7.7%×40%) to 68.3% (i.e., 60% + 20.8%×40%). Although this predicted overall increase is relatively small it does not seem to be seen in Fig. 12 for any of the AGE and “Alpha variant” adjusted *IDE* versions.

Next, we move on to the period with the greatest daily testing rate examined, which is the beginning of May 2021. At that time about 3% of the Danish population was PCR tested on a daily basis. In addition to this, about twice that number was tested daily using Rapid Antigen tests. However, as previously noted, RA tests only have a sensitivity of about 50% of PCR tests for asymptomatic cases. Thus, we assume that effectively ∼6% of the Danish population is equivalently PCR tested on a daily basis (i.e., 3% PCR + 6% RA×50% effective PCR). When correcting for the 10-day positivity period and compensating for multiple detection of positive individuals during those 10 days (assuming purely random testing), 46.1% of the asymptomatic cases should be caught by the large-scale testing. If that value is multiplied by the 40% of cases that we had assumed were asymptomatic, then the value of the

*IDE* would increase from the 60% value due to symptomatic cases to 78.4% (i.e., 60% + 46.1%×40%) in total, an overall increase of 18.4 percentage points. Averaging out the semi-cyclic fluctuations this is about the increase seen in Fig. 12 for the AGE and “Alpha variant” adjusted *IDE* using the Danish hospitalization adjustment. The increase seen for the AGE and “Alpha variant” adjusted *IDE* using the Austrian hospitalization adjustment is slightly less, being about 15% as seen in Fig. 12. Finally, for the AGE and “Alpha variant” adjusted *IDE* using the ≥ 60 Simple Method, virtually no increase is seen, which completely contradicts the predictions.

On the other hand, there are a series of issues with the values seen in Fig. 12 for all the AGE and “Alpha variant” adjusted *IDE* values for 2021. First, for the first few months of 2021 the *IDE* values are less than their 2020 “*steady*” period values by at least 5 percentage points. Having seen that the “*steady*” period values were essentially associated with symptomatic cases and also “*normal*” testing/contact tracing efforts, the *IDE* values should not decline unless testing notably drops. However, as seen in the upper plot of Fig. 14, during the period of low *IDE* values in 2021 the per person testing rate for Denmark as a whole is comparable if not greater than the rate during the “*steady*” period, especially relative to the early part of the “*steady*” period. Overall, the drop in *IDE* values would seem to imply that during the first few months of 2021 the testing effort is not as effective as in 2020. In fact, it is almost as if the targeting is aimed at the non-infected part of the population and/or that symptomatic individuals are no longer always coming in to be tested. While this is unexpected, it somehow seems to be the case. With regard to expected behavior, the HOPE-Project found that by late 2020 and continuing into early 2021 the Danish population that had been in close contact with infected individuals did not get tested at nearly the level one would hope for and furthermore, only ∼50% of the close contact population obeyed self-isolation instructions [48, 49]. The ∼50% fraction was also on a downward trend entering 2021 with a similar trend possibly occurring for symptomatic individuals coming in to be tested.

Another issue is the semi-cyclical peak and valley behavior of the *IDE* from about mid-March 2021 onwards. The behavior is all the more surprising as the values plotted in Fig. 12 are 7-day running average values. In looking for a cause of the peaks, when the timing of the peaks in Fig. 12 is compared with the per person PCR testing rate and/or the PCR test positivity values, no correlations are seen. [The per person RA testing rate (not shown) also has no correlation with the semi-cyclical *IDE* peaks.] Thus, we have another example of unexpected and strange impacts of the testing effort in 2021 on the *IDE* values. This is indicative of a less-than-ideal implementation of the testing effort to be able to catch asymptomatic cases. Another work [50] also found poor performance of Denmark’s large-scale testing effort at its height in 2021.

### 4.3 COVID-19 mitigation performance: Denmark vs. other countries

It is of interest to compare the effectiveness of Denmark’s large contact tracing and testing program in terms of per capita COVID cases, deaths, and tests per detected COVID case to the same quantities for selected other countries and regions. These values from Ref. [14] are shown in Fig. 15 for 4 different time periods. The June 16 – Oct. 15, 2020 values span a time when the number of COVID19 cases were relatively low in many countries due to the summer conditions. Also, during that period, most northern hemisphere countries were still expanding their testing capability and seemingly achieved a capacity that was more than sufficient to meet demand during that period. This conclusion was inferred by examining the ratio of the number of tests performed per detected case as a function of time that could be calculated from values from Ref. [14]. The ratio values typically rose from small values early in the pandemic when testing capacity was insufficient and then began to plateau at the start of summer and remain relatively constant throughout the summer while the number of new COVID-19 infections tended to decline while the testing capacity continued to increase for most countries. However, as the number of infections began to increase during the transition to winter, the ratio of tests to the number of detected COVID-19 cases decreased indicative of testing capacity once again not meeting demand. This was also true for Denmark.

**Fig. 15.**
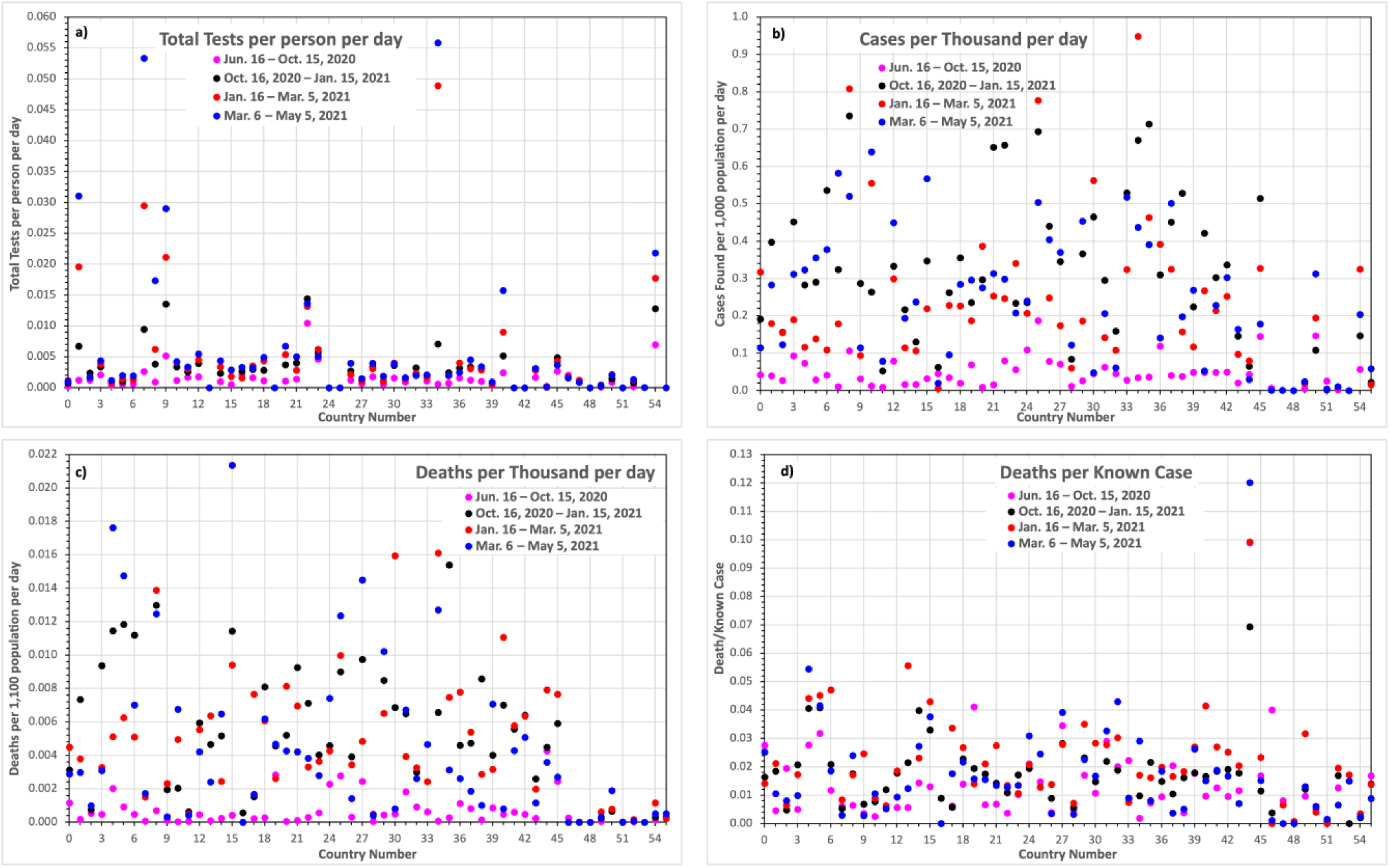
As a function of “country” the plots show: a) The total number of COVID tests done per person per day; *b) The total number of COVID cases found per 1,000 people per day; c) The total number of COVID deaths found per 1,000 people per day”; d) The total number of COVID deaths found per COVID case”. Values are given for 4 different time periods. The “countries” are as follows: 0:Albania, 1:Austria, 2:Belarus, 3:Belgium, 4:Bosnia - Herzegovina, 5:Bulgaria, 6:Croatia, 7:Cyprus, 8:Czechia, 9:Denmark, 10:Estonia, 11:Finland, 12:France, 13:Germany, 14:Greece, 15:Hungary, 16:Iceland, 17:Ireland, 18:Italy, 19:Kosovo, 20:Latvia, 21:Lithuania, 22:Luxembourg, 23:Malta, 24:Moldova, 25:Montenegro, 26:Netherlands, 27:North Macedonia, 28:Norway, 29:Poland, 30:Portugal, 31:Romania, 32:Russia, 33:Serbia, 34:Slovakia, 35:Slovenia, 36:Spain, 37:Sweden, 38:Switzerland, 39:Ukraine, 40:United Kingdom, 41:Europe, 42:European Union, 43:Canada, 44:Mexico, 45:United States, 46:Australia, 47:New Zealand, 48:China, 49:Japan, 50:Kuwait, 51:Singapore, 52:South Korea, 53:Taiwan, 54:United Arab Emirates, 55:Asia. The values are from Ref. [14] and the tests values don’t differentiate between antigen and PCR varieties. Note that several countries did not report the number of tests given (e.g. China) and that test values are also not reported for continents or regions (e.g., Asia, Europe, or European Union)*.

In examining the values shown in Fig. 15, it should be noted that test values are not reported for several countries, nor are they reported for regions. The reported number of tests include antigen tests in addition to PCR tests. Furthermore, when comparing the values it is important to take into account external factors that can significantly modify values of one country versus another. For instance, the number of cases and deaths for both Australia and New Zealand are very low simply because being “island” nations that were able enforce strict isolation policies as opposed to extensive testing of the population as a whole. Another example is Mexico, which had a very high number of deaths per known cases simply because it did relatively little testing and hence there were very low number of “known” cases relative to other countries.

In comparing the values for Denmark (Country 9) it is seen that it had among the highest number of tests per person per day, a relatively low number of cases per capita per day, and a very low number of COVID deaths per capita per day. While the high number of tests per capita done in Denmark no doubt contributed to this desirable outcome, it must be noted that the ∼5 tests per person from the pandemic start through May 5, 2021 (which corresponds to ∼1 test per person in 3 months) is still quite small and insufficient to prevent the spread of the pandemic.

As a contrast to Denmark, and most other countries depicted in Fig. 15, Sweden (Country 37) did not impose any national or local lockdowns and also didn’t impose restrictive measures such as closing preschools and elementary schools. However, measures were taken to protect the most vulnerable and there was a ban on large gatherings as well as guidelines for social distancing. The Swedish citizens were mainly asked to follow voluntary ‘principles of responsibility’ [51]. Initially, this meant significantly higher infection and death rates compared to Denmark, but surprisingly, integrated over the pandemic 2020-2022 the estimated excess mortality rates for Denmark and Sweden both turned out to be quite low ∼80 deaths/100k vs. ∼117 deaths/100k respectively [52]. This means that there are alternative ways to successfully weather a pandemic. It should nonetheless be noted that the stricter measures of Denmark did achieve a COVID-19 death rate that was just under 70% (i.e. 80/117) of that of Sweden.

### 4.4 Costs associated with COVID-19 containment efforts

Danmarks Statistik (Statistics Denmark – the Danish authority on Danish statistics) estimated the COVID-19 health related costs for 2020 and 2021 [53]. They found that the total health expenses for Denmark in 2021 were about 270.8 billion Danish Krone (270.8 B DKK or ∼36.4 B Euro) or about 10.8% of Denmark’s GNP. This was 25.5 B DKK more than in 2020. For 2020, the COVID-19 direct health care related expenses were 5.1 B DKK of which 1.7 B DKK were for contact tracing and testing efforts. In contrast, the same costs for 2021 were respectively 20.1 B DKK and 12.5 B DKK.

Danmarks Statistik also reported values for the overall COVID-19 costs for all of 2020 plus the first 2 quarters of 2021 [54]. That value was 80.1 B DKK overall, which was distributed as follows: 18.5 B DKK (23.1%) for direct COVID-19 health care related expenses; 56.9 B DKK (71.0%) for financial compensation; and the bulk of the remaining 4.7 B DKK (5.9%) going to targeted initiatives for both specific industries and the social areas. Many more costs could be included in such an analysis, most notably costs due of the loss of economic productivity. Also note that the positive value of fewer lives lost and a more open society due to better preparedness are not included in the values given. However, for our purpose, a simple order of magnitude estimate based on the above set of values is sufficient to make the following arguments.

Aside from masks, lockdowns and other societal restrictions, the combination of contact tracing and testing preparedness is one of the primary defenses against the spread of the pandemic before the availability of vaccines. The main goal of these initiatives was to prevent an overwhelming of the healthcare system, which was successful. However, as noted in Section 4.2, the testing and contact tracing program had to be more focused and robust than it was on average to control the spread of the pandemic, as virtually all asymptomatic infected were not identified. From the values above it is seen that only 1.7 B DKK [of the 18.5 B DKK spent on testing and contact tracing from the start of the pandemic through Quarter 2 (Q2) of 2021] was spent in all of 2020. It should also be noted that the total amount spent on COVID-19 healthcare was about 1/3 of that spent on financial compensation for direct COVID-19 economic losses through Q2 of 2021. Furthermore, the amount spent on testing and contact tracing in 2020 was only ∼3% of the financial compensation.

While there are startup times associated with building up a testing and contact tracing program, the small amount spent in 2020 on contact tracing and testing was not of a scale needed to control the pandemic aside from the instance when it was highly focused in Aarhus, where it was also combined with a local lockdown. The time frame when a high level of buildup was needed was in the summer of 2020 and before the fall and winter period when respiratory diseases typically rapidly spread. To partly answer our previous question “How much preparedness is enough?” it seems that too little (only 3% of the total direct public expenditure costs) was invested too late.

As an increasing number of infected weren’t detected and isolated in the fall and early winter of 2020, a large outbreak occurred that necessitated a second countrywide lockdown with large financial compensation expenditures.

### 4.5 The presented *IDE* method and its limitations

The presented *IDE* method is based on national/regional antibody prevalence measurements together with daily hospital admission data which form the foundation for our ability to estimate an *Infection Detection Efficiency* (*IDE*) on a daily basis; see Sections 2 and 3. Daily hospital admission data stemming from the pandemic can be used to track the actual size of the pandemic including asymptomatic cases when the prevalence number is known. To do this, a variety of adjustments from other data sources must be applied to the hospital admission numbers, most importantly COVID variant and AGE-based corrections, the latter of which can be obtained in different ways. Which way is adopted gives slightly different results, see Sections 3.6, 3.7, and 3.8. The adjusted hospital admission values are summed and scaled to match the size of the pandemic available from the antibody prevalence measurements. Even with reliable antibody tests, population wide prevalence data are both difficult to obtain and inherently associated with uncertainty, which at best leads to an estimated ‘true’ *Infection Detection Efficiency*. For example, using endpoints of the 95% CI around the selected prevalence value used herein from sections 2.3 and 3.3, an estimated average *IDE* value of 60.0% spans the range between 56.76 % and 63.64%. Despite these limitations it is our belief that an implementation of the *IDE* provides a critical daily measure of how a society is mitigating a pandemic.

## 5. Conclusions and Lessons Learned

We have changed our approach for investigating the actual scale of the COVID-19 pandemic in Denmark from one based on simulations [3] to one relying on published data. We also expanded the time frame covered from that of our earlier work [3] and now cover the period from the start of the pandemic into May 2021, a time when vaccines were already playing a noticeable role with about 1/4 of the Danish population having had received at least one COVID-19 vaccine dose. The main emphasis of the work herein was an evaluation of the ability to find all of Denmark’s COVID-19 cases through testing and contact tracing and from that determining the ability of such efforts to be used as a means of containing and controlling the pandemic.

From the work detailed herein a list of lessons learned that should be applied to future pandemics follows:

1. National/regional antibody prevalence measurements together with daily hospital admission data are the foundation of our ability to estimate the actual scale of the COVID-19 pandemic and to determine an *Infection Detection Efficiency* (*IDE*) on a daily basis; see Sections 2 and 3.
2. Daily hospital admission data stemming from the pandemic can be used to track the actual size of the pandemic including asymptomatic cases when the prevalence number is known. To do this, a variety of adjustments from other data sources must be applied to the hospital admission numbers, see Section 3. The adjusted hospital admission values are summed and scaled to match the measured integral size of the pandemic available from the antibody prevalence measurements. Due to the many intricacies in these estimations and to speed up availability of reliable pictures of the pandemic, an open data and method exchange between research groups are preferred including governmentally assigned pandemic study teams as well as the open international academic community.
3. Influenza-like airborne viral infections are more common in the winter months, with less sunlight, lower temperatures and humidity, and far more indoor activities, compared to the summer months [e.g. 44–47]. Thus, a relatively quiet summer period should be used to invest and prepare both civil and governmental initiatives for an upcoming winter period.
4. A difficult balance between restrictive policies and free choices of citizens must be achieved, which is an open question of how best to do, especially as it is likely to be country dependent and possibly even region dependent within a country. The authors of this paper do not envy the policymakers for having to strike such a balance. Obviously, policies that restrict gatherings, as well as nationwide targeted mandatory testing and contact tracing, together with mandatory quarantine, are in principle more effective in pandemic control than voluntary actions, but such strict mandatory policy actions are very difficult to implement in an open democratic society such as Denmark.

From our analysis of the Danish COVID-19 pandemic we can emphasize:

5) The daily number of new COVID-19 cases derived as above were then compared with the number of COVID-19 cases confirmed in real time via PCR testing. It was found that the combined PCR testing/contact tracing program as “*normally*” employed does not find ∼ 40% of COVID cases that are likely asymptomatic according to the literature; namely (100% – *IDE*) ≍ Asymptomatic case fraction; see Section 4.
6) However, when contact tracing and testing are employed in an enhanced and targeted manor as was done briefly in Aarhus, essentially all of the local COVID cases were found and the localized outbreak was brought under control; see Section 4.2. Also, the essentially 100% detection efficiency achieved in that one instance indicates that in principle added restrictions and lockdowns can be avoided when enhanced, well-targeted testing and contact tracing are implemented. Unfortunately, the testing and contact tracing effort as “*normally*” employed did not work as a means controlling the pandemic.
7) The capacity of the testing program met demand by the summer of 2020 in part because the number of Denmark’s COVID-19 cases was low due to the March 2020 shutdown and the summer conditions that were far less conducive to respiratory disease spreading as noted in point 3). Unfortunately, this led to a false sense of security. There was a strong wish to believe that the pandemic was under control, which was also communicated by the Danish government. This in turn led to a failure to make the investments needed to significantly expand the contact tracing and testing capacities in preparation for the serious late 2020 pandemic growth that should have been foreseen. Also, the lax social gathering policies that made sense in the summer were not updated to consider the impact of back-to-school gatherings and colder weather with more indoor activities. Although the contact tracing and testing capacity continued to be slowly expanded in the summer–early–fall period, it could not catch up with the rapid rise in cases in the fall and late 2020 [11,12]. This eventually necessitated a costly national lockdown Dec. 25, 2020, to bring the outbreak under control; see Section 4.
8) From July 2020 through early May 2021, only 2 significant peaks in the *IDE* were detected. Unlike the Aarhus peak mention in point 6) above, the mid-Nov. 2020 peak could not be attributed to any specific changes in governmental policy and/or changes in testing and contact tracing activities. In particular, examining the testing efforts in the different regions of Denmark during that time, contrary to popular associations, the mid-Nov. 2020 *IDE* peak was shown NOT to be related to North Jutland efforts connected to the mink outbreak.
9) Attempts to catch a larger fraction of cases using higher levels of “*normal*” testing and contact tracing was minimally successful at best. Furthermore, the extensive use of RA tests was far less helpful than predicted due to their poor performance for asymptomatic cases, which were exactly the cases that needed to be caught to contain the pandemic.
10) The drop of the *IDE* in the first few months of 2021 to values only slightly over 50%, which is less than the normal fraction of symptomatic cases implied that a noticeable fraction of the Danish population was no longer responding as desired or expected when ill and displaying COVID-like symptoms. This assertion was bolstered by the findings mentioned earlier [49] that as of late 2020 and early 2021 only about 50% of the Danish population was obeying isolation requirements.
11) Lastly, we want to mention a problem we, the authors, experienced as scientists from the open academic community working on the Danish part of the pandemic. We suggest this problem should be resolved for a future pandemic:

i. As mentioned earlier, ALL relevant key data collected and/or generated by the government agencies should be made publicly available to the open research community. While most data (e.g., observed daily infected, hospitalizations, ICU, deaths, prevalence, …) was indeed available, this was not the case for all of the important data (e.g., the daily contact tracing activities). Obviously, all issues regarding data privacy should be respected for the published data, and governmentally collected and compiled data cannot be expected to be immediately available in the first chaotic month of the pandemic. However, longer term lack of data access hampers the ability of the open scientific community to help develop new insights of potentially critical value for public health and policy decisions.
ii. To harness the rapid COVID-19 knowledge development more fully from the open international scientific community, the responsible Danish governmental agencies should have allowed a proper and more direct knowledge and data sharing between the working expert groups within the governmental agencies and the external open scientific community. This would have allowed the internal expert groups to request external input to pertinent national health and policy issues as well as second opinions and on-the-fly peer support/review. Further, new pertinent insights could have been transferred directly from the outside to the inside, without first having to be published through the traditional scientific channels. In a small country as Denmark, weekly or biweekly, moderated, telecom meetings could have been established. In such meetings the internal expert groups could have requested input to pertinent scientific issues, while external groups could have presented pertinent new results and insights. The telecom moderation could have been done by a small group of experts both from the inside and from the outside, once the first chaotic month of the pandemic was over and proper working routines were established. Immediately after the onset of the pandemic this online mode of scientific collaboration and exchange became well-established throughout the open international scientific community. As an example of cross-agency collaboration in the US, researchers from the Santa Fe Institute (from the open academic community) collaborated via telecoms with staff from the National Institute of Health (NIH) and the U.S. Centers for Disease Control and Prevention (CDC), which are both central official US governmental agencies mandated by the US Government to develop pandemic mitigations. A similar beneficial collaborative effort between the government and the open scientific community could have been established in Denmark.

Despite the above discussed issues, Denmark did very well compared to most other countries in terms of limiting deaths and preventing hospital overflows. This was likely due to Denmark’s relatively large contact tracing and testing program and its government-imposed restrictions on interactions. The overarching National Danish pandemic priority was to ensure the healthcare systems were not overwhelmed by the pandemic, while keeping society as open as possible. In the big picture this balance was achieved in part through the implemented restrictions, a large testing effort, and both formal and informal infection tracing efforts combined with changed citizens’ behaviors. However, the testing and contact tracing program as “*normally*” implemented could not do it alone and unfortunately nationwide lockdowns were required twice to bring the pandemic back under control. In only one instance for Denmark were enhanced testing and contact tracing efforts employed well enough to bring outbreaks under control on a large scale. Unfortunately, these efforts were not maintained long enough or widely enough to prevent the pandemic from reestablishing itself.

## Data Availability

All data produced in the present study are available upon reasonable request to the authors. The starting data sources are all referenced in the paper.

## 6. Acknowledgement

We are extremely grateful for the help of Michael Skytte Liboriussen whose work on our previous studies [3] and contributions to this work led to the current paper.

## Notes

### Competing Interest Statement

The authors have declared no competing interest.

### Author Declarations

With one exception, all the data used in this paper are "published", publicly available, and are referenced in the paper. The only exception is some purely statistical values that were the result of private communications which are also referenced in the paper, (Reference [16]).

